# Genome-wide SNP-sex interaction analysis of susceptibility to idiopathic pulmonary fibrosis

**DOI:** 10.1101/2024.01.12.24301204

**Authors:** Olivia C Leavy, Anne F Goemans, Amy D Stockwell, Richard J Allen, Beatriz Guillen-Guio, Tamara Hernandez-Beeftink, Ayodeji Adegunsoye, Helen L Booth, CleanUP-IPF Investigators of the Pulmonary Trials Cooperative, Paul Cullinan, William A Fahy, Tasha E Fingerlin, Harvinder S Virk, Ian P Hall, Simon P Hart, Mike R Hill, Nik Hirani, Richard B Hubbard, Naftali Kaminski, Shwu-Fan Ma, Robin J McAnulty, X Rebecca Sheng, Ann B Millar, Maria Molina-Molina, Vidya Navaratnam, Margaret Neighbors, Helen Parfrey, Gauri Saini, Ian Sayers, Mary E Strek, Martin D Tobin, Moira KB Whyte, Yingze Zhang, Toby M Maher, Philip L Molyneaux, Justin M Oldham, Brian L Yaspan, Carlos Flores, Fernando Martinez, Carl J Reynolds, David A Schwartz, Imre Noth, R Gisli Jenkins, Louise V Wain

## Abstract

**Background:** Idiopathic pulmonary fibrosis (IPF) is a chronic lung condition that is more prevalent in males than females. The reasons for this are not fully understood, with differing environmental exposures due to historically sex-biased occupations, or diagnostic bias, being possible explanations. To date, over 20 independent genetic variants have been identified to be associated with IPF susceptibility, but these have been discovered when combining males and females. Our aim was to test for the presence of sex-specific associations with IPF susceptibility and assess whether there is a need to consider sex-specific effects when evaluating genetic risk in clinical prediction models for IPF.

**Methods:** We performed genome-wide single nucleotide polymorphism (SNP)-by-sex interaction studies of IPF risk in six independent IPF case-control studies and combined them using inverse-variance weighted fixed effect meta-analysis. In total, 4,561 cases (1,280 females and 2,281 males) and 23,500 controls (8,360 females and 14,528 males) of European genetic ancestry were analysed. We used polygenic risk scores (PRS) to assess differences in genetic risk prediction between males and females.

**Findings:** Three independent genetic association signals were identified. All showed a consistent direction of effect across all individual IPF studies and an opposite direction of effect in IPF susceptibility between females and males. None had been previously identified in IPF susceptibility genome-wide association studies (GWAS). The predictive accuracy of the PRSs were similar between males and females, regardless of whether using combined or sex-specific GWAS results.

**Interpretation:** We prioritised three genetic variants whose effect on IPF risk may be modified by sex, however these require further study. We found no evidence that the predictive accuracy of common SNP-based PRSs varies significantly between males and females.

**Research in context:** 

**Evidence before this study:** The prevalence of IPF is higher in males than females. IPF risk has a genetic component, but analyses have only been performed in studies where males and females have been combined. One previous study reported sex-specific differences in association for the *MUC5B* promoter variant, rs35705950, however the finding was not replicated in an independent study. No genome-wide association studies assessing for different genetic risk factors between males and females have been conducted for IPF. It is not known whether approaches to predict individuals at risk of IPF should take sex-specific genetic risk into consideration.

**Added value of this study:** This was the largest study to test whether there are genetic variants whose effects on IPF susceptibility are different in males and females. The *MUC5B* promotor variant rs35705950 did not show a different magnitude of effect in males vs females. We identified three genetic variants with opposite directions of effect on IPF risk in males vs females. Our polygenic risk score analyses suggested that genetic prediction based on data from males and females separately did not perform better than when males and females were combined.

**Implications of all available evidence:** Although we found some preliminary evidence of genetic variants with sex-specific effects on IPF risk, our analyses suggest that genome-wide genetic risk from common single nucleotide polymorphisms is similar in males and females. This is important when considering integration of polygenic risk scores into clinical prediction models for IPF. There may be other forms of genetic variation, such as complex structural variation or rare variants, not captured in this analysis, that may improve risk prediction for males and females separately.

## Introduction

Idiopathic pulmonary fibrosis (IPF) is a progressively fibrotic lung disease with a median survival time after diagnosis of 3-5 years^1^. In the USA and Europe, IPF is estimated to have a disease prevalence of 0.63 to 7.6 per 100,000 people^2^. The number of people diagnosed with IPF is increasing and males are more likely to be diagnosed than women^3,4^. However, the reason why the disease is more prevalent in males is not understood. Different environmental exposures between males and females, notably occupations such as carpentry which have traditionally been more common amongst men^5^, could explain some of the observed difference. Diagnostic bias may also play a role with men being over diagnosed and women being undiagnosed with IPF^6^. As well as prevalence differences, there are survival differences with men having worse survival after IPF diagnosis than females^7^. Differences in genetic predisposition between males and females may be an additional factor in prevalence differences, however, this has not yet been extensively studied.

IPF is a complex polygenic disease with multiple genes implicated in susceptibility. The genetic variant rs35705950 in the *MUC5B* gene promoter has been shown to increase a person’s risk of IPF 5-fold for each copy of the risk allele^8–11^ and has been estimated to explain more than three times more disease liability than the other known common IPF risk variants combined^12^. In recent years, genome-wide association studies (GWAS), examining genetic variants across the genome, have identified over 20 genetic loci associated with IPF risk^13–16^. In addition to providing new insight into disease biology, polygenic risk scores (PRS) derived from GWAS data have shown potential utility in identifying individuals at highest risk of pulmonary fibrosis^17^.

We hypothesised that there might be different biological mechanisms that promote IPF susceptibility in males and females, and that genetic associations that differ between males and females, may pinpoint the genes and pathways involved. To test this, we performed a genome-wide single nucleotide polymorphism (SNP)-by-sex interaction meta-analysis of IPF risk in six independent clinically-defined IPF case-control studies. Given the increasing interest in the use of PRS as a clinical tool for diagnosis in complex diseases^18^, we additionally tested whether PRS derived from sex-combined association data performed differently in males and females or whether derivation of PRS from sex-specific data might improve predictive accuracy.

## Methods

### Studies

We performed a SNP-by-sex interaction meta-analysis for IPF risk using six independent IPF case-control studies, all of which have been previously described; US^19^ (formerly referred to as Chicago), Colorado^13^, UK^20^, UUS^11^, Genentech^21^ and CleanUP-UCD^22,23^ **(Table 1)**. In short, unrelated participants from the six studies were included in this analysis if they were of genetically-determined European ancestry and had sex-at-birth recorded. We only included participants who passed genotyping quality control and cases were defined using the relevant American Thoracic Society/European Respiratory Society guidelines^24,25^ (**Supplementary Methods**).

**Table 1:**
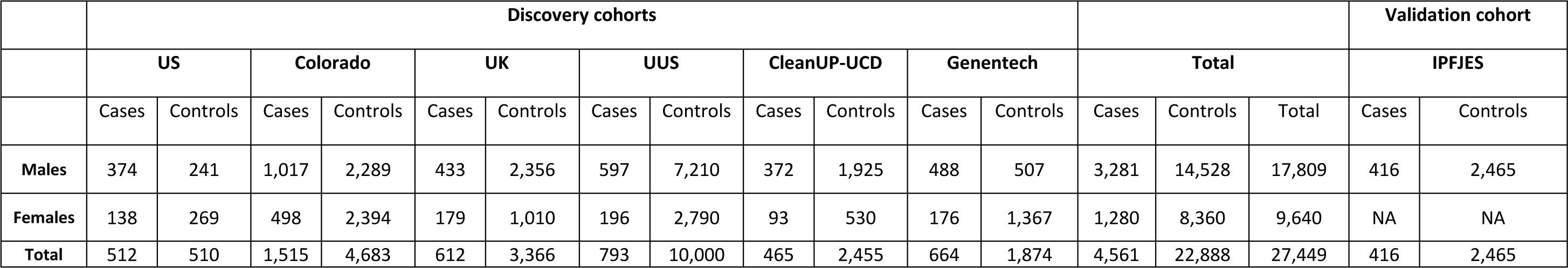
IPF case-control cohorts.

### Genome-wide SNP-by-sex Interaction Analyses

Genome-wide SNP-by-sex interaction analyses of IPF risk were performed separately in each of the six studies and meta-analysed using PLINK 1.9 (www.cog-genomics.org/plink/1.9/)^26^ (**Supplementary Methods**). Analyses were performed using autosomal SNPs. We used *P*<5x10^-8^ as the threshold for genome-wide significance and *P*<1x10^-6^ for suggestive significance in the meta-analysis. Independent sentinel variants were defined using distance-based and conditional analysis methods (**Supplementary Methods**).

As all available datasets with both male and female participants were included within the genome-wide discovery analysis to maximise statistical power, we applied Meta-Analysis Model-based Assessment of replicability (MAMBA^27^) to assess the posterior probability of replication for all SNPs with a meta-analysis *P*<1×10^−6^. SNPs with a MAMBA posterior probability of replication (PPR) >90% were considered to be robust across the contributing studies and likely to replicate in future studies.

Male-specific and female-specific effect estimates were calculated for all sentinel variants passing the above criteria. We sought validation of male-specific effect sizes and direction in a male-only IPF case control study, IPFJES (IPF Job Exposure Study)^28^, comprising 416 male IPF cases and 2,465 male controls (**Figure 1**, **Supplementary Methods**). No independent female-only datasets were available at the time of conducting this study.

**Figure 1:**
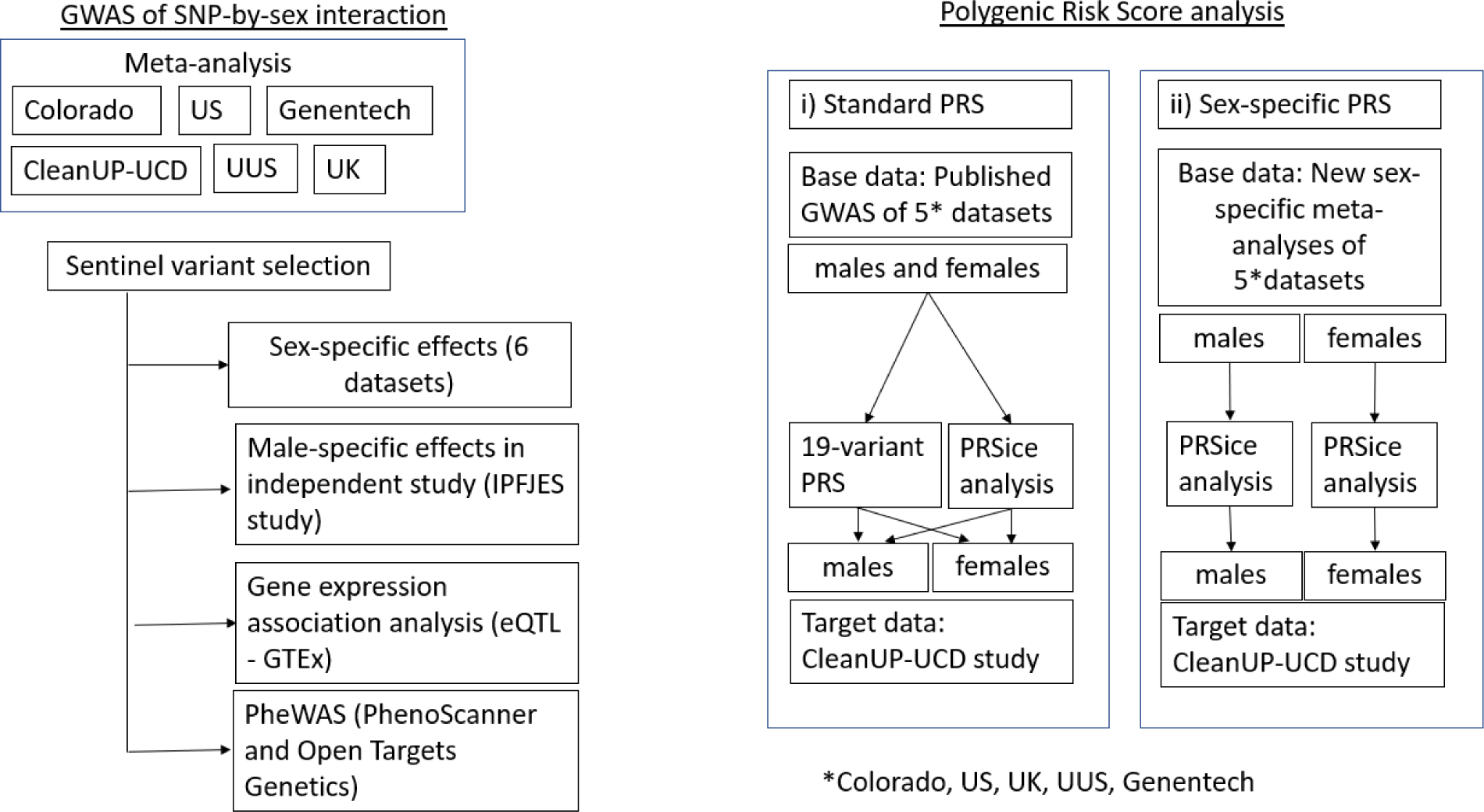
Overview of the SNP-by-sex interaction analysis and polygenic risk score analysis

### Bioinformatic investigation of significant signals

Annotation of variants was performed using Variant Effect Predictor (VEP)^29^. We used GTEx to assess whether the sentinel variants were eQTLs for gene expression in up to 49 tissues (including lung and non-lung tissues) and the coloc package^30^ in R version 4.2.1 to test if sentinel variants were eQTLs for gene expression in lung or cultured fibroblasts (**Figure 1, Supplementary Methods**). We conducted phenome-wide association studies (PheWAS) using PhenoScanner^31,32^ and Open Targets^33^ to examine whether the signals were also associated with other phenotypes.

### Polygenic risk score analyses

As well as looking at the effect individual genetic variants have on disease risk, the effect multiple SNPs have can be explored using PRS. The scores are constructed by taking the weighted sum (usually weighted by the SNP effects from a GWAS data) across many SNPs. It can then be tested whether these scores are predictors of disease risk. We wanted to test whether the predictive accuracy of PRS in predicting IPF risk differed between males and females. The predictive accuracy of PRS was evaluated in two ways: 1) the predictive performance of the PRS derived from sex-combined IPF susceptibility GWAS^14^ was evaluated in males and females separately (‘standard PRS’) and 2) the predictive performance of the PRS derived from sex-specific GWAS was evaluated in males and females separately (‘sex-specific PRS’) (**Figure 1**). For 1) we first evaluated a 19-variant PRS representing previously reported common genome-wide significant (*P*<5x10^-8^) signals of association with IPF^14^. For both 1) and 2) we further incorporated genome-wide data using an approach that varied the threshold used for inclusion of variants in the PRS (PRSice v2.3.5^34^). ‘Base data’ were derived from sex-combined and sex-specific meta-analyses of the US, Colorado, UK, UUS and Genentech datasets. The ‘target dataset’ was the independent CleanUP-UCD study comprising 465 cases (93 females and 372 males) and 2,455 controls (530 females and 1,925 males). Area Under the Curve (AUC) differences were tested using DeLong’s test (**Supplementary Methods**).

## Results

The genome-wide sex interaction analysis of IPF risk was performed in up to 4,561 cases (comprising 1,280 females and 2,281 males) and 23,500 controls (8,360 females and 14,528 males). A total of 8,485,642 genetic variants were included in the meta-analysis and there was no evidence of inflated test statistics **(Figure 2 & Figure S1)**.

**Figure 2:**
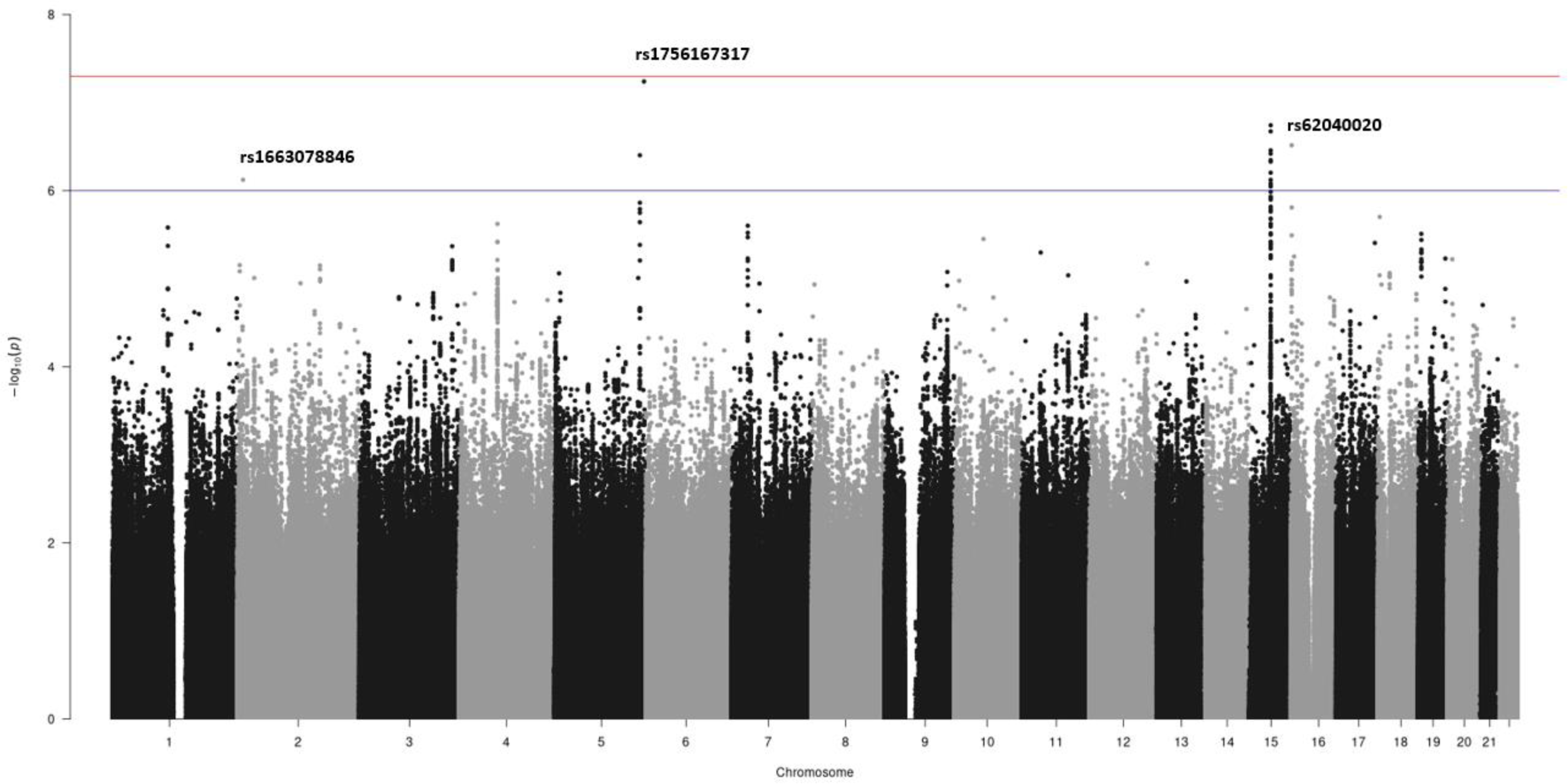
Manhattan plot of meta-analysed sex-interaction results. The chromosomal position is on the x-axis and the -log(*p*-value) for each genetic variant in the sex-interaction meta-analysis is on the y-axis. Variants present in at least 3 studies are presented. The blue horizontal line represents the 1×10^−6^ *p*- value threshold and the red horizontal line represents 5×10^−8^ *p*-value threshold (genome-wide significance threshold).

Three independent sentinel variants with interaction *P*<1×10^−6^ and MAMBA PPR>90% were identified **(Table S1)**. All three variants had consistent direction of effect across all contributing studies **(Figure 3a, b, c)**.

**Figure 3:**
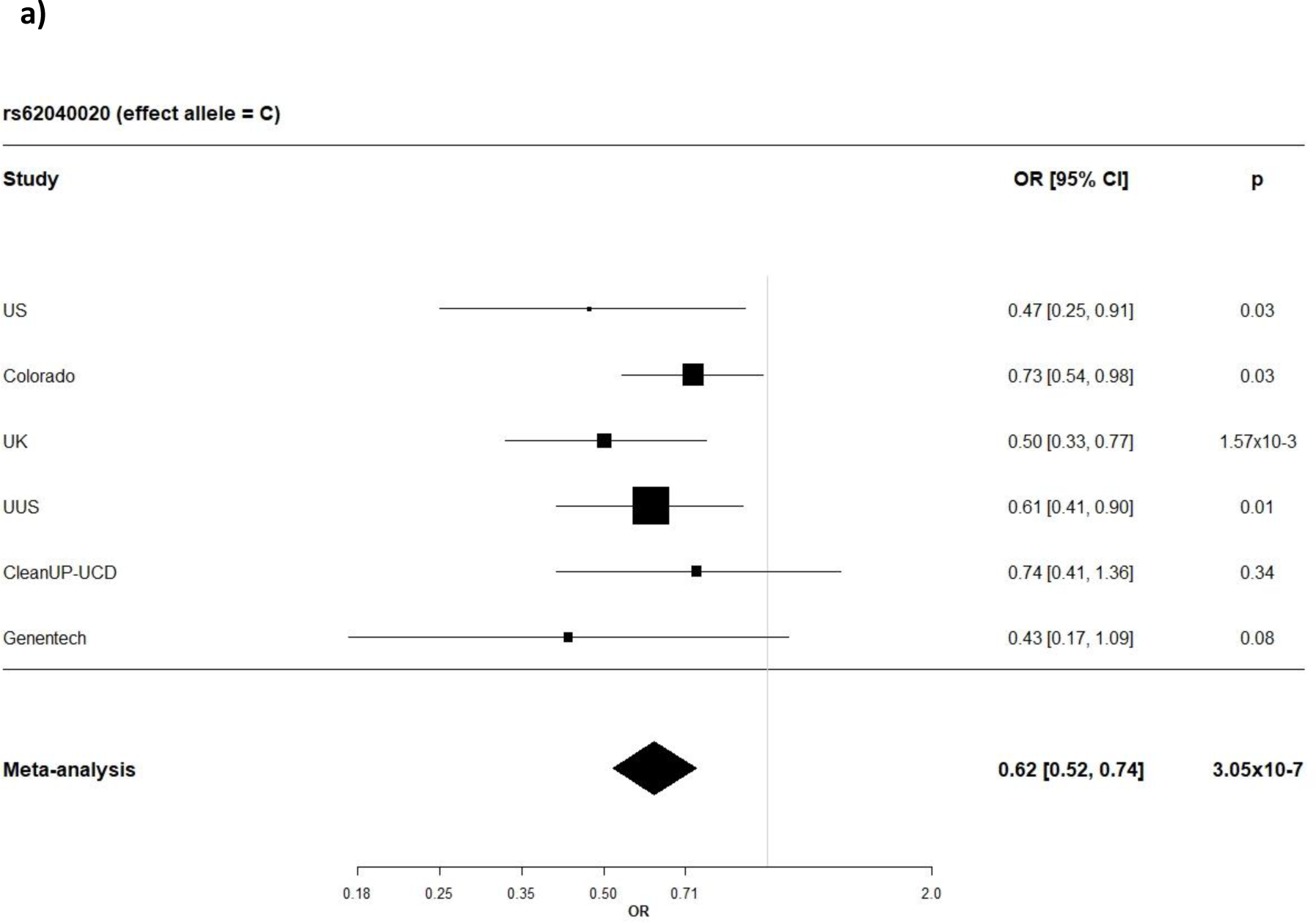
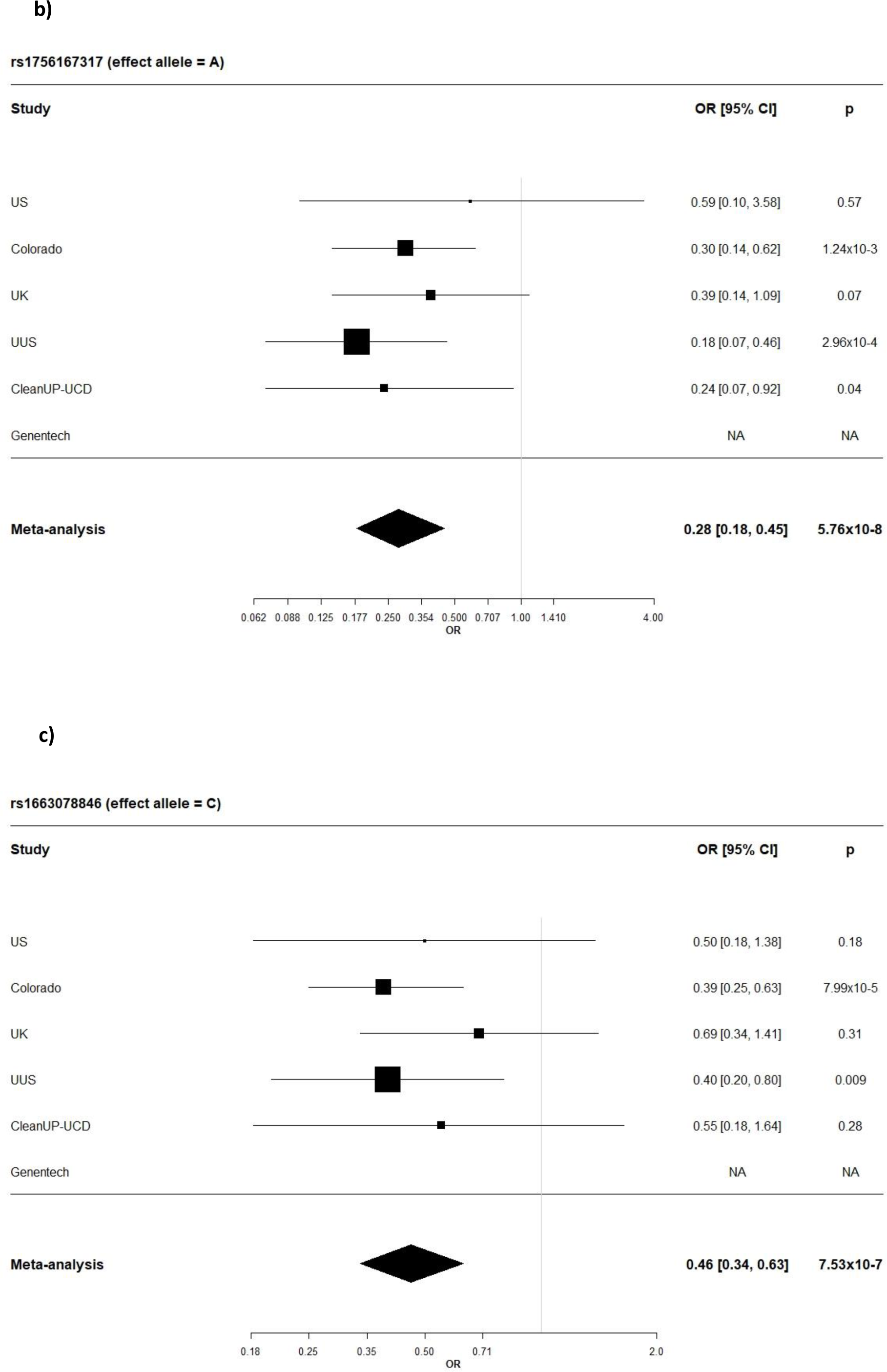
Forest plots showing SNP-sex interaction odds ratio by study and the meta-analysed results for **a)** rs62040020, **b)** rs1756167317 and **c)** rs1663078846. OR = odds ratio and CI = confidence interval

The sentinel variant rs62040020, which resided within an intron of *JPT2* (Jupiter microtubule associated homolog 2) on chromosome 16 (effect allele frequency (EAF) = 10.6%), was measured in all six studies with a high imputation quality (R^2^ > 0.88 across all six studies) and was nominally significant (*P*<0.05) in four of the six studies **(Table S1 & Figure 4a)**. When tested for association with IPF risk in females and males separately, the minor allele (allele = C) of rs62040020 was associated with increased risk of IPF in females (odds ratio (OR) 1.34, 95% confidence intervals (CI) 1.15-1.55, *P*=1x10^-4^) and decreased risk in males (OR 0.82, 95% CI 0.74-0.92, *P*=1x10^-3^) **(Figure 5)**. Accordingly, if we instead took the major allele to be the effect allele (allele = G), the direction of effect would be in the opposite direction (i.e., increased risk of IPF in males and decreased risk in females). In the male-only IPFJES study, the association effect was close to the null and non-significant (OR 0.99, 95% CI 0.77-1.27, *P*=0.939) **(Figure S2a)**. In lung and/or cultured fibroblasts, the C allele of rs62040020 (associated with increased IPF risk in females, decreased risk in males) was associated with increased expression of *FAHD1* (Fumarylacetoacetate Hydrolase Domain Containing 1), *MEIOB* (meiosis specific with OB-fold) and *NUBP2* (NUBP Iron-Sulfur Cluster Assembly Factor 2, Cytosolic) and decreased expression of *MRPS34* (mitochondrial ribosomal protein S34) **(Table S2)**. This allele was also related to changes in splicing of *HAGH* (hydroxyacylglutathione hydrolase) in lung, cultured fibroblasts and a range of other tissues. However, rs62040020 was not the most significant variant associated with expression of these genes at this locus, with colocalisation analyses for male sex-specific GWAS results suggesting the GWAS and eQTL association signals were driven by different variants (**Figure S3 & Table S3, Supplementary Methods**). The C allele of this variant was previously associated with reduced monocyte percentage in UK Biobank (NEALE round 2 results: http://www.nealelab.is/uk-biobank/, *P*=1.7x10^-4^) **(Table S2)**.

**Figure 4:**
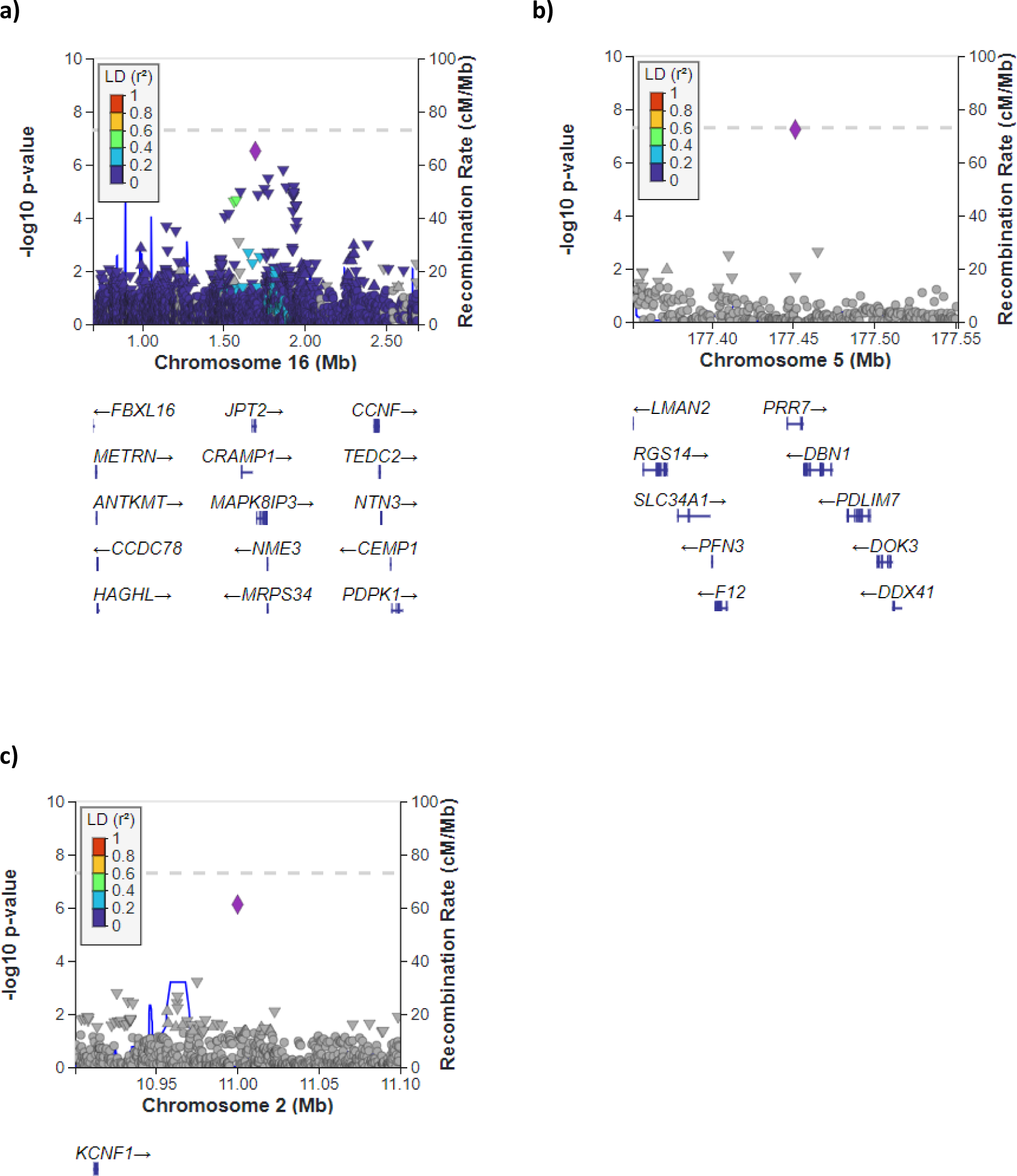
Region plots for **a)** rs62040020, **b)** rs1756167317 and **c)** rs1663078846. The chromosomal position is on the x-axis and the -log(*p*-value) for each genetic variant in the sex- interaction meta-analysis is on the y-axis.

**Figure 5:**
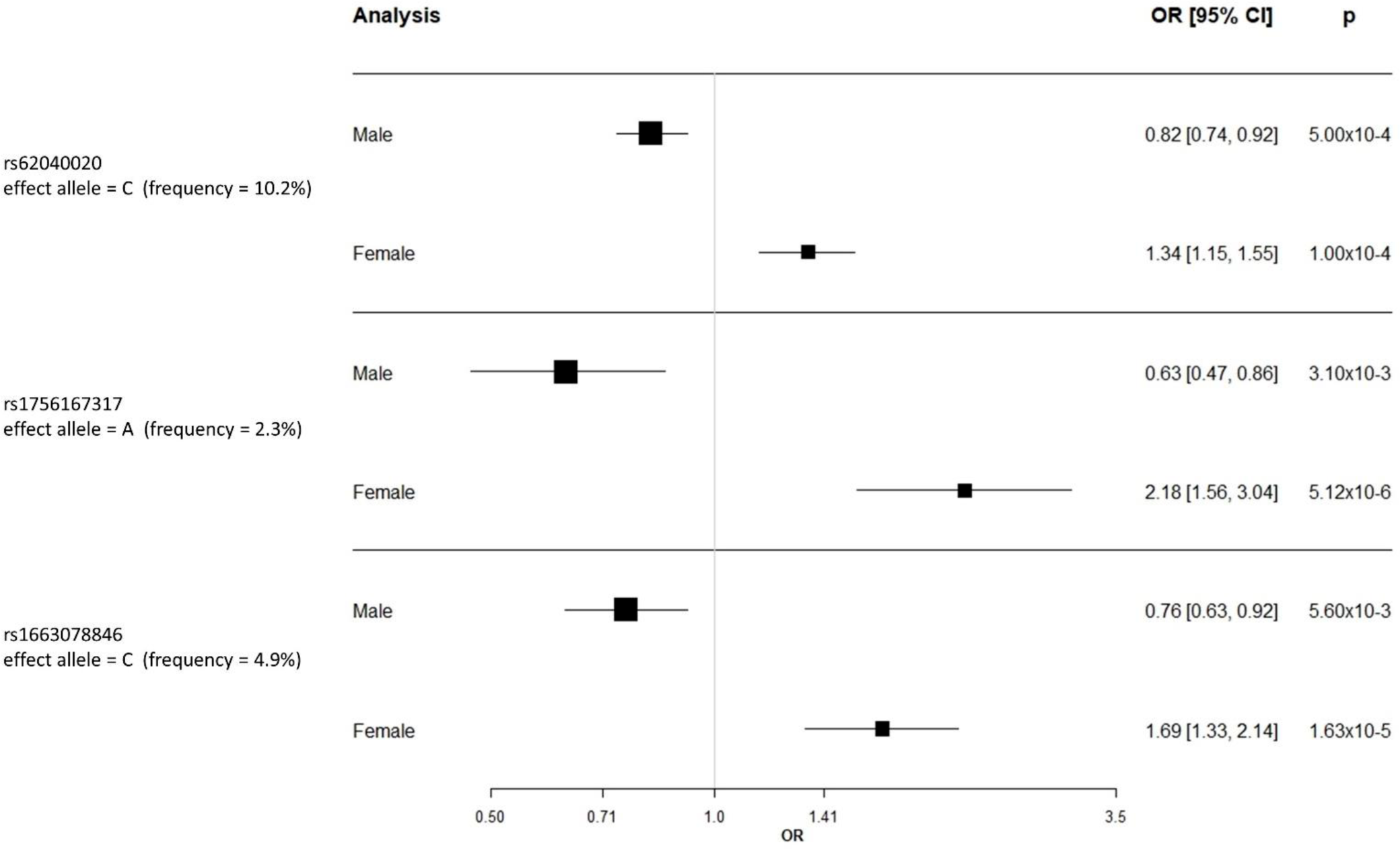
Forest plot for sex stratified results meta-analysed across US, Colorado, UK, UUS, CleanUP- UCD and Genentech. OR = odds ratio and CI = confidence interval

For the other two SNPs, rs1756167317 and rs1663078846, the sentinel variants were of low frequency (EAF: 1-5%) and were nominally significant in 3 out of 5 contributing studies and 2 out of 5 contributing studies, respectively (**Figures 3b, 3c & Table S1)**. The A allele of rs1756167317, located in an intron of *PRR7* (Proline rich 7, synaptic) at chromosome 5 (**Figure 4b)**, was associated with increased risk of IPF in females (OR 2.18, 95% CI 1.56-3.04, *P*=5x10^-6^) and decreased risk in males (OR 0.63, 95% CI 0.47-0.86, *P*=3x10^-3^) **(Figure 5)**. This SNP showed a consistent direction and size of effect in the male only IPFJES study (OR 0.68, 95% CI 0.36-1.30, *P*=0.247) **(Figure S2b)**. For the sentinel variant of the signal on chromosome 2, rs1663078846, the C allele was associated with increased risk of IPF in females (OR 1.69, 95% CI 1.33-2.14, *P*=2x10^-5^) and decreased risk in males (OR 0.76, CI 0.63-0.92, *P*=6x10^-3^) **(Figure 5)**. This intergenic variant (**Figure 4c**) did not show a consistent effect in males in the IPFJES study (OR 1.07, 95% CI 0.74-1.55, *P*=0.702)**(Figure S2c)**. In PhenoScanner and Open Targets no associations with other traits for rs1756167317 or rs1663078846 were found.

No variants met genome-wide significance (*P*<5×10^−8^), one was borderline genome-wide significant (rs1756167317, *P* = 5.76x10^-8^). None of the common previously reported IPF susceptibility variants, including the *MUC5B* promoter variant rs35705950, were observed to have a significant sex interaction effect when accounting for multiple testing **(Table S4)** although the *DSP* (desmoplakin) variant, rs2076295, had a nominally significant interaction effect (*P*=0.03) (**Figure S4**).

### Polygenic risk scores

For the ‘standard PRS’ analysis, there was no difference in the AUC between males and females for the 19-variant PRS (AUC males: 80.3% vs AUC females: 80.8%, DeLong *P* = 0.85) **(Table S5)**. When constructing multiple PRS (i.e., not limiting to published IPF susceptibility variants) the most predictive *p*-value threshold (*P*T ) was *P*T<4.5×10^-4^ for males and *P*T<5×10^-4^ for females **(Figure S5a & Figure S5b)**, and whilst the AUC estimated was slightly lower for males than for females, the difference was not statistically significant (male AUC: 80.2% vs female AUC: 81.8%, DeLong *P* = 0.54).

For the ‘sex-specific PRS’ analysis, the male specific PRS predictive accuracy was slightly higher than the female specific PRS predictive accuracy, but the difference was not statistically significant (male-specific PRS AUC: 78.2% vs female-specific PRS AUC: 76%, DeLong *P* = 0.47) **(Table S5) (Figure S5c & Figure S5d)**. The AUCs observed in this analysis were smaller than those generated in the ‘standard PRS’ analysis, which might be explained by the decrease in sample size of the training set (i.e., less accurate effect sizes).

The PRS results suggest that the predictive accuracy of IPF PRSs is not statistically different between males and females, regardless whether using combined or sex-specific GWAS results.

## Discussion

We performed the first genome-wide SNP-by-sex interaction analysis of IPF risk in clinically-defined cases and identified three independent signals that were suggestively significant at *P*<1×10^−6^. To test whether a combination of variant effects (including known IPF susceptibility variants, as well as those not reaching statistical significance) predict IPF susceptibility in males and females differently, we performed PRS analyses. We found that the predictive accuracies of PRSs were not sex-dependent suggesting that PRS developed from sex-combined association statistics are largely generalisable across sexes.

Although none of the genetic variants analysed reached genome-wide significance in our interaction analysis, three sentinel variants met a less stringent threshold for significance and were consistent across all studies included in the meta-analysis. None of the variants replicated in the male-only IPFJES study, but this could have been because the study was under-powered to validate the male-specific effects. Further data are needed to provide confidence in these signals and to confirm the likely causal genes at these loci. These new signals however may offer further insight into sex-specific mechanisms in IPF. *FAHD1*, *HAGH* and *MRPS34* have all been implicated in mitochondrial function; mitochondrial dysfunction has been widely implicated in age-related disease such as IPF^35^. *HAGH* was also amongst 2,940 genes differentially expressed between IPF cases and controls^36^.

However, the sex-specific effect of these genes has not been investigated.

None of the 19 previously reported common IPF genetic variants^14^ demonstrated a suggestively significant sex-interaction (*P* < 1×10^−6^). Previously, a sex-stratified meta-analysis conducted across six biobank studies^15^ observed a larger effect size in males compared with females for the *MUC5B* variant rs35705950. The effect was not replicated in a clinically-defined IPF sub-study of FinnGen or in the four clinically-defined case-control studies (four of the six studies used were included in our present study). We also did not observe this difference in our analysis (which included additional datasets). We have previously highlighted differences in genetic association effect sizes when defining IPF from routine electronic healthcare records compared to clinically defined cohorts^37^ suggesting that case definition heterogeneity might account for the biobank finding.

Studies of interaction effects require larger sample sizes than GWAS of main effects on disease risk as we are testing for a difference in effect size between two subgroups of participants; we cannot exclude the possibility that there are additional sex-specific genetic association signals yet to be discovered with larger sample sizes. However, our PRS analysis also included variants not meeting stringent statistical significance and did not suggest that there were any large effect sex-specific signals yet to be detected. There were more males than females in the analysis, which would affect the 95% CI of the AUC, with females having a wider AUC 95% CI than males.

It could be that there are sex differences, but we might not see these differences on the autosomal chromosomes as sex chromosomes were not analysed. Furthermore, given the known role of mitochondrial function it may be that sex differences might be observed in mitochondrial DNA. Our PRS analysis was not intended to define the optimum PRS for IPF in either or both sexes, but rather to indicate whether future efforts should focus on sex-specific PRS development. Our study was limited to variants with MAF>1% and as such we cannot exclude the potential for rare variants with sex-specific effects. All data included in this study was derived from individuals of European ancestry; our findings may not be generalisable to other ancestries and larger genetic studies of ILD in non-European ancestry populations, with appropriate representation of both sexes, are urgently needed.

In summary, our genome-wide SNP-by-sex interaction analysis identified three potential sex-interaction signals which require further validation and functional investigation. Our polygenic risk score analysis suggests that PRS derived from sex-combined IPF SNP association studies perform similarly in males and females with no significant benefit in deriving sex-specific PRS.

## Data Availability

Full summary statistics for genome-wide SNP-sex interaction meta-analyse can be accessed from https://github.com/genomicsITER/PFgenetics.

## Ethics

This research was conducted with appropriate ethics approval.

The PROFILE study (which provided samples for the UK and UUS studies) had institutional ethics approval at the University of Nottingham (NCT01134822 – ethics reference 10/H0402/2) and Royal Brompton and Harefield NHS Foundation Trust (NCT01110694 – ethics reference 10/H0720/12). UK samples were recruited across multiple sites with individual ethics approval (University of Edinburgh Research Ethics Committee [The Edinburgh Lung Fibrosis Molecular Endotyping (ELFMEN) Study NCT04016181] 17/ES/0075, NRES Committee South West – Southmead, Yorkshire and Humber Research Ethics Committee 08/H1304/54 and Nottingham Research Ethics Committee 09/H0403/59). Spanish samples were recruited under ethics approval by ethics committee from the Hospital Universitario N.S. de Candelaria (reference of the approval: PI-19/12). The UUS study also included individuals from clinical trials with ethics approval (ACE [NCT00957242] and PANTHER [NCT00650091]). For the UCSF cohort, sample and data collection were approved by the University of California San Francisco Committee on Human Research and all patients provided written informed consent. For the Vanderbilt cohort, the Institutional Review Boards from Vanderbilt University approved the study and all participants provided written informed consent before enrolment. For individuals recruited at the University of Chicago, consenting patients with IPF who were prospectively enrolled in the institutional review board-approved ILD registry (IRB#14163A) were included. Individuals recruited at the University of Pittsburgh Medical Centre had ethics approval from the University of Pittsburgh Human Research Protection Office (referenceSTUDY20030223: Genetic Polymorphisms in IPF). Individuals from the COMET (NCT01071707) and Lung Tissue Research Consortium (NCT02988388) studies were also included in the Chicago study. All subjects in the Colorado study gave written informed consent as part of IRB-approved protocols for their recruitment at each site and the GWAS study was approved by the National Jewish Health IRB and Colorado Combined Institutional Review Boards (COMIRB). Subjects in the Genentech study provided written informed consent for whole-genome sequencing of their DNA. Ethical approval was provided as per the original clinical trials (INSPIRE [NCT00075998], RIFF [NCT01872689], CAPACITY [NCT00287729 and NCT00287716] and ASCEND [NCT01366209]).

Individuals in the CleanUP-UCD study included individuals from clinical trials with ethics approval (NCT02759120). These samples were genotyped under University of Virginia ethics approval (IRB 20845). IPFJES involved human participants and was approved by East Midlands - Nottingham 1 Research Ethics Committee REC reference: 17/EM/0021IRAS project ID: 203355. Participants gave informed consent to participate in the study before taking part.

## Funding and Acknowledgements

RGJ and LVW report funding from the Medical Research Council (MR/V00235X/1). LVW holds a GSK / Asthma + Lung UK Chair in Respiratory Research (C17-1). The research was partially supported by the National Institute for Health Research (NIHR) Leicester Biomedical Research Centre and the NIHR Imperial Biomedical Research Centre; the views expressed are those of the author(s) and not necessarily those of the National Health Service (NHS), the NIHR or the Department of Health. RGJ is funded by an NIHR Research Professorship (RP-2017-08-ST2-014). BGG is supported by Wellcome Trust grant 221680/Z/20/Z. NK reports funding from National Institutes of Health (R01HL127349) and is a scientific founder at Thyron, served as a consultant to Boehringer Ingelheim, Pliant, Astra Zeneca, RohBar, Veracyte, Augmanity, CSL R01HL141852, U01HL145567, R21HL161723, P01HL11450, (NK). CF is supported by Ministerio de Ciencia e Innovación RTC-2017-6471-1 (AEI/FEDER UE), Instituto de Salud Carlos III PI20/00876 cofinanced by the European Regional Development Fund “A Way of Making Europe’’ from the European Union; Instituto Tecnologico y de Energias Renovables agreement OA23/043. JMO funding from the NIH (NHLBI) (R01HL169166).

The UK, UUS CleanUP-UCD and IPFJES studies selected controls from UK Biobank under application 648. This research used the SPECTRE High Performance Computing Facility at the University of Leicester. For the purpose of open access, a CC BY or equivalent licence will be applied to any author accepted.

## GTEx Portal

The Genotype-Tissue Expression (GTEx) Project was supported by the Common Fund of the Office of the Director of the National Institutes of Health, and by NCI, NHGRI, NHLBI, NIDA, NIMH, and NINDS. The data used for the analyses described in this manuscript were obtained from: the GTEx Portal on 19/05/22.

## Global Biobank Engine

Global Biobank Engine, Stanford, CA (URL: http://gbe.stanford.edu) [19^th^ May 2022 accessed]. G. McInnes, Y. Tanigawa, C. DeBoever, A. Lavertu, J. E. Olivieri, M. Aguirre, M. A. Rivas, Global Biobank Engine: enabling genotype-phenotype browsing for biobank summary statistics. Bioinformatics (2019). doi:10.1093/bioinformatics/bty999

## Conflicts of Interest

AA declares funding from NIH (K23HL146942); consulting fees from Genentech, Inogen, Medscape, Abbvie, PatientMpower and Boehringer Ingelheim; payment or honoraria for lectures, presentations, speakers bureaus, manuscript writing or educational events from Boehringer Ingelheim. ADS is a full-time employee of Genentech/Roche with stock and stock options in Roche. AFG was a full-time employee of PPD, Part of Thermo Fisher Scientific until June 2023. BGG declares fellowship funding from Wellcome Trust (221680/Z/20/Z). BLY is a full-time employee of Genentech/Roche with stock and stock options in Roche. CF declares funding Ministerio de Ciencia e Innovación, Instituto de Salud Carlos III and Instituto Tecnológico y de Energías Renovables; honoraria in educational events from Fundación Instituto Roche. DAS declares being the founder and chief scientific officer of Eleven P15, Inc., a company dedicated to the early diagnosis and treatment of pulmonary fibrosis. HP declares grant payment to institution from Boehringer Ingelheim Ltd; consulting fees from Boehringer Ingelheim Ltd, Roche Limited, Trevi Therapeutics, Pilant Therapeutics; speaker fees from Boehringer Ingelheim Ltd; member of TIPAL trial management group, trustee for Action for Pulmonary Fibrosis, member of scientific advisory board for European Pulmonary Fibrosis Federation. IN declares funding from National Institutes of Health (UG3HL145266) to institution; grant funding to institution from Veracyte; consulting fees from Boerhinger Ingelheim and Sanofi. IPH declares funding from Wellcome Trust and NIHR; vice chair Trustees for Asthma + Lung UK. JMO declares funding from National Institutes of Health (R01HL169166 & K23HL138190); consulting fees from Boehringer Ingelheim, Lupin pharmaceuticals, AmMax Bio, Roche and Veracyte; patent for TOLLIP TT genotype for NAC use in IPF; participation on a Data Safety Monitoring Board or Advisory Board for Endeavor Biomedicines, Novartis and Genentech; Associate editor for CHEST, on Program Committee for American Thoracic Society and Editorial board for AJRCCM. LVW declares funding from UK Research and Innovation (MR/V00235X/1) and GSK/Asthma + Lung UK (Professorship (C17-1)) to complete this work; funding from Orion Pharma, GSK, Genentech, AstraZeneca, Nordic Bioscience, Sysmex (OGT); Consulting fees Galapagos, Boehringer Ingelheim, GSK; support for attending meetings and/or travel Genentech; participation on Advisory Board for Galapagos; leadership or fiduciary roles as Associate Editor for European Respiratory Journal and Medical Research Council Board member and Deputy Chair.

MKBW declares funding from National Institutes of Health (K23HL146942); consulting fees from Genentech, Inogen, Medscape, Abbvie, PatientMpower and Boehringer Ingelheim; payment or honoraria for lectures, presentations, speakers bureaus, manuscript writing or educational events from Boehringer Ingelheim. MN is a full-time employee of Genentech/Roche with stock and stock options in Roche. NK declares grant funding from National Institutes of Health; grant funding to institution from BMS, Boehringer Ingelheim and Three Lakes Foundation; consultancy fees from Biogen Idec, Boehringer Ingelheim, Third Rock, Pliant, Samumed, NuMedii, Theravance, Three Lake Partners, Astra Zeneca, RohBar, Veracyte, Augmanity, CSL Behring, Thyron, Gilead, Galapagos, Chiesi, Arrowhead, Sofinnova, GSK and Merk; patent for new therapies for IPF (Biotech), new therapies for ARDS (Biotech) and new Biomarkers in IPF (Biotech); equity in Pilant and Thyron; reports serving as the scientific founder of Thyron. PLM declares grant funding to institution from AstraZeneca; consultancy fees from Hoffman-La Roche, Boehringer Ingelheim, AstraZeneca, Trevi and Qureight; speaker fees from Boehringer Ingelheim and Hoffman-La Roche. RGJ declares funding from UK Research and Innovation (MR/V00235X/1); that their institute received funding from Astra Zeneca, Biogen, Galecto, GlaxoSmithKline, Nordic Biosciences, RedX and Pliant; consulting fees from AstraZeneca, Brainomix, Bristol Myers Squibb, Chiesi, Cohbar, Daewoong, GlaxoSmithKline, Veracyte, Resolution Therapeutics and Pliant; payment for lectures and presentations received from Boehringer Ingelheim, Chiesi, Roche, PatientMPower, AstraZeneca; payment for expert testimony from Pinsent Masons LLP; participation on a Data Safety Monitoring Board or Advisory Board for Boehringer Ingelheim, Galapagos, Vicore; leadership or fiduciary role for NuMedii and president for Action for Pulmonary Fibrosis. SPH declares grant funding to institution from Boehringer Ingelheim; consulting fees from Trevi therapeutics; payment or honoraria for lectures, presentations, speakers bureaus, manuscript writing or educational events from Chiesi and Trevi therapeutics; support for attending meetings and/or travel from Chiesi; Participation on a Data Safety Monitoring Board or Advisory Board for Trevi therapeutics; Chair for BTS Standards of Care Committee (till November 2022) and Trustee for Action for Pulmonary Fibrosis. TMM declares consulting fees from Boehringer Ingelheim, Roche/Genentech, Astra Zeneca, Bayer, Blade Therapeutics, Bristol-Myers Squibb, CSL Behring, Galapagos, Galecto, GlaxoSmithKline, IQVIA, Pfizer, Pliant, Respivant, Sanofi, Theravance, Trevi, Veracyte and Vicore; participation on a Data Safety Monitoring Board or Advisory Board for Fibrogen, Blade Therapeutics and Nerre. XRS is a full-time employee of Genentech/Roche with stock and stock options in Roche.

## Notes

### Author Declarations

This research was conducted with appropriate ethics approval. The PROFILE study (which provided samples for the UK and UUS studies) had institutional ethics approval at the University of Nottingham (NCT01134822 - ethics reference 10/H0402/2) and Royal Brompton and Harefield NHS Foundation Trust (NCT01110694 - ethics reference 10/H0720/12). UK samples were recruited across multiple sites with individual ethics approval (University of Edinburgh Research Ethics Committee [The Edinburgh Lung Fibrosis Molecular Endotyping (ELFMEN) Study NCT04016181] 17/ES/0075, NRES Committee South West - Southmead, Yorkshire and Humber Research Ethics Committee 08/H1304/54 and Nottingham Research Ethics Committee 09/H0403/59). Spanish samples were recruited under ethics approval by ethics committee from the Hospital Universitario N.S. de Candelaria (reference of the approval: PI-19/12). The UUS study also included individuals from clinical trials with ethics approval (ACE [NCT00957242] and PANTHER [NCT00650091]). For the UCSF cohort, sample and data collection were approved by the University of California San Francisco Committee on Human Research and all patients provided written informed consent. For the Vanderbilt cohort, the Institutional Review Boards from Vanderbilt University approved the study and all participants provided written informed consent before enrolment. For individuals recruited at the University of Chicago, consenting patients with IPF who were prospectively enrolled in the institutional review board-approved ILD registry (IRB#14163A) were included. Individuals recruited at the University of Pittsburgh Medical Centre had ethics approval from the University of Pittsburgh Human Research Protection Office (referenceSTUDY20030223: Genetic Polymorphisms in IPF). Individuals from the COMET (NCT01071707) and Lung Tissue Research Consortium (NCT02988388) studies were also included in the Chicago study. All subjects in the Colorado study gave written informed consent as part of IRB-approved protocols for their recruitment at each site and the GWAS study was approved by the National Jewish Health IRB and Colorado Combined Institutional Review Boards (COMIRB). Subjects in the Genentech study provided written informed consent for whole-genome sequencing of their DNA. Ethical approval was provided as per the original clinical trials (INSPIRE [NCT00075998], RIFF [NCT01872689], CAPACITY [NCT00287729 and NCT00287716] and ASCEND [NCT01366209]). Individuals in the CleanUP-UCD study included individuals from clinical trials with ethics approval (NCT02759120). These samples were genotyped under University of Virginia ethics approval (IRB 20845). IPFJES involved human participants and was approved by East Midlands - Nottingham 1 Research Ethics Committee REC reference: 17/EM/0021IRAS project ID: 203355. Participants gave informed consent to participate in the study before taking part.

### Summary of Updates

Changes to the order of the authorship

