## Supplementary Methods for "Genome-wide SNP-sex interaction analysis of susceptibility to idiopathic pulmonary fibrosis"

[1]Department of Population Health Sciences, University of Leicester, Leicester, UK, [2]NIHR Leicester Biomedical Research Centre, Leicester, UK, [3]Genentech, California, USA, [4]University of Chicago, Chicago, USA, [5]University College London Hospitals, London, UK, [6]Weill Cornell Medicine, New York, USA, [7]Imperial College, London, UK, [8]GlaxoSmithKline, London, UK, [9]National Jewish Health, Colorado, USA, [10]University of Nottingham, Nottingham, UK, [11]National Institute for Health Research, Nottingham Biomedical Research Centre, Nottingham, UK, [12]University of Hull, Hull, UK, [13]University of Oxford, Oxford, UK, [14]University of Edinburgh, Edinburgh, UK, [15]Yale School of Medicine, Connecticut, USA, [16]University of Virginia, Virginia, USA, [17]University College London, London, UK, [18]University of Bristol, Bristol, UK, [19]Servei de Pneumologia, Laboratori de Pneumologia Experimental, Instituto de Investigación Biomédica de Bellvitge (IDIBELL), Barcelona, Spain, [20]Campus de Bellvitge, Universitat de Barcelona, Barcelona, Spain, [21]Centro de Investigación Biomédica en Red de Enfermedades Respiratorias (CIBERES), Instituto de Salud Carlos III, Madrid, Spain, [22]Department of Respiratory Medicine, Sir Charles Gardiner Hospital, Perth, Australia, [23]Centre for Respiratory Research, University of Western Australia, Perth, Australia, [24]Royal Papworth Hospital NHS Foundation Trust, Cambridge, UK, [25]Centre for Respiratory Research, NIHR Nottingham Biomedical Research Centre, School of Medicine, Biodiscovery Institute, University of Nottingham, Nottingham, UK, [26]University of Pittsburgh, Pittsburgh, USA, [27]NIHR Imperial Biomedical Research Unit, National Heart and Lung Institute, Imperial College London, London, UK, [28]Division of Pulmonary and Critical Care Medicine, University of Southern California, Los Angeles, USA, [29]National Institute for Health Research Respiratory Clinical Research Facility, Royal Brompton Hospital, London, UK, [30]University of Michigan, Michigan, USA, [31]Research Unit, Hospital Universitario Nuestra Señora de Candelaria, Santa Cruz de Tenerife, Spain, [32]Genomics Division, Instituto Tecnologico y de Energias Renovables, Santa Cruz de Tenerife, Spain, [33]Facultad de Ciencias de la Salud, Universidad Fernando Pessoa Canarias, Las Palmas de Gran Canaria, Spain, [34]University of Colorado Medicine, Colorado, USA

#### International IPF Genetics Consortium

**UK Study**
Richard J Allen, Helen L Booth, William A Fahy, Ian P Hall, Simon P Hart, Mike R Hill, Nik Hirani, Richard B Hubbard, R Gisli Jenkins, Toby M Maher, Robin J McAnulty, Ann B Millar, Philip L Molyneaux, Vidya Navaratnam, Eunice Oballa, Helen Parfrey, Gauri Saini, Ian Sayers, Martin D Tobin, Louise V Wain, Moira K B Whyte

**US Study**
Ayodeji Adegunsoye, Carlos Flores, Naftali Kaminski, Shwu-Fan Ma, Imre Noth, Justin M Oldham, Mary E Strek, Yingze Zhang

**Colorado Study**
Tasha E Fingerlin, David A Schwartz

**UUS Study**
Richard J Allen, Carlos Flores, Beatriz Guillen-Guio, R Gisli Jenkins, Shwu-Fan Ma, Toby M Maher, Maria Molina-Molina, Philip L Molyneaux, Imre Noth, Justin M Oldham, Louise V Wain

**Genentech study**
Margaret Neighbors, X Rebecca Sheng, Amy Stockwell, Brian L Yaspan

**CleanUP-UCD**
Fernando Martinez, Imre Noth, CleanUP-IPF Investigators of the Pulmonary Trials Cooperative

**IPFJES**
Paul Cullinan, Carl Reynolds

### Supplementary Methods

#### Overview of study

In this study we have used six IPF case-control studies (US, Colorado, UK, UUS, Genentech and CleanUP-UCD) to conduct a genome-wide SNP-by-sex interaction meta-analysis.

Interaction analyses were performed in each of the six studies separately, using genetic variants imputed using the Trans-Omics for Precision Medicine (TOPMed) reference panel and the Michigan Imputation Server[1]. The individual study results were then meta-analysed using a fixed-effect meta-analysis in order to improve the statistical power of the analysis. From the meta-analysis we used meta-$P < 1x10^{-8}$ as the threshold for genome-wide significance and the threshold meta-$P < 1\times10^{-6}$ was used to define suggestively significant interactions. The IPF study IPFJES was used to assess males only results from the meta-analysis.

We performed polygenic risk score (PRS) analyses with the scores constructed for CleanUP-UCD males and female participants separately, using the SNP effects from meta-analysing five of the IPF case-control cohorts. We used the area under the ROC curve (AUC) and DeLong's test to test whether there were differences in the predictive accuracy of multiple PRS between males and females.

#### Methods

##### Studies and quality control

Quality control (QC) and sample selection for five of the six studies meta-analysed (US, Colorado, UK, UUS, Genentech) has previously been described in Allen *et al*[2] and the CleanUP-UCD study[3, 4] has been described separately.

In the analysis we retained IPF cases and controls who were of genetically-determined European ancestry and had sex-at-birth recorded.

IPFJES (the IPF Job Exposure Study) has been described by Reynolds *et al*[5]. The study comprises 960 (494 cases and 466 controls) individuals who were recruited across 21 UK hospitals. IPF cases were male who had been first diagnosed with IPF between 1st February 2017 and 1st October 2019. Controls were selected from non-IPF individuals attending outpatient's departments over the same time-period.
Of the 494 IPF cases and 466 controls, 441 cases and 423 controls were genotyped by the Affymetrix UK Biobank array.

The following subject quality control was performed:

- Affymetrix quality control: four individuals removed after failing Affymetrix dish quality control and five individuals removed as they had a call rate < 97% in step 1 genotype calling (performed using Axiom Power Tools).
- Individual call rate: four individuals removed as they had an individual call rate < 95%.
- Sex mismatches: two individuals removed as inferred genetic sex using PLINK 1.9 (www.cog-genomics.org/plink/1.9/)[6] was different to the recorded sex.
- Duplicates and relatedness: relatedness was estimated using KING[7] on autosomal genetic variants with a genotype call rate > 95%, minor allele frequency (MAF) > 1%, in Hardy-

Weinberg equilibrium ($P > 10^{-6}$) and not found to be in regions of high linkage disequilibrium. Duplicate/monozygotic twin pairs were defined as those with kingship coefficient > 0.3540, first-degree relatives were those with a kingship coefficient between 0.1770 and 0.3540 and second-degree relatives were those with a kingship coefficient between 0.0884 and 0.1770. Seven individuals identified as being duplicate/monozygotic twins were removed and 13 individuals were removed due to high relatedness (first-degree or second-degree relatedness). One individual was removed as they had already been genotyped in other IPF cohort studies (UK and UUS studies).

- Ancestry: principal component analysis, using PLINK on the genetic data and HapMap samples, was used to infer ancestry. Autosomal genetic variants that were present in HapMap with a genotype call rate > 95%, MAF > 1%, in Hardy-Weinberg equilibrium ($P > 10^{-6}$) and not found to be in regions of high linkage disequilibrium were used in the principal component analysis. In total, 27 individuals were identified as being of non-European ancestry.

After subject quality control, 416 IPF cases and 385 controls were retained. Additional controls from UK Biobank were combined with the 385 controls to improve statistical power. The number was increased to 2,465 controls so that there were 5 controls to every case (additional 2,080 UK Biobank controls were selected). Imputation was performed using the TOPMed imputation reference panel.

#### Genome-wide SNP*sex interaction analysis

Genome-wide sex interaction analyses of IPF risk were performed separately in each study, using PLINK 1.9.

The following logistic regression model was applied:

$$\text{Phenotype}_i = \beta_0 + \beta_1 \text{G}_i + \beta_2 \text{Sex}_i + \beta_3 \text{G}_i \text{Sex}_i + \text{PC}_{1i} + \ldots + \text{PC}_{10i} + \varepsilon_i$$

where,

*Phenotype* is IPF status, *G* is the dosage for a given genotype (additive effect), *Sex* is binary coded, *G*Sex* is the interaction term and $PC_1$ to $PC_{10}$ are the first ten standardised genetic principal components for ancestry.

Bi-allelic autosomal variants that were well imputed (imputation $R^2 \geq 0.5$) using the TOPMed imputation reference panel, had a MAF ≥ 0.01, did not depart from Hardy-Weinberg Equilibrium ($P \geq 1\times10^{-6}$) and were present in at least three of the six studies were retained in the analysis. The results from each of the six IPF cohorts were then combined using an inverse-variance weighted fixed effect meta-analysis, implemented using PLINK 1.9. Genomic control lambda was calculated for the meta-analysis.

We defined sentinel variants as those with the smallest *p*-value and $P<1\times10^{-6}$ within a 1 Mb region. Then GCTA-COJO[8] was used to identify additional independent signals meeting $P<1\times10^{-6}$ within each 1 Mb window.

Region plots were produced using LocusZoom[9].

#### Bioinformatic investigation of signals

Fine-mapping was used to produce a set of genetic variants that had a 95% probability of containing the causal variant for a given genetic signal (95% credible set). This was performed in R version 4.2.1.

The fine-mapping approach used (Wakefield approximate Bayes factor) assumed that there was one causal variant and that it had been measured.

We used GTEx Portal to check if the genetic variants in the 95% credible sets were associated with gene expression across 49 tissues (including lung and non-lung tissues). For those found to be associated with expression levels of a gene in either lung or cultured fibroblasts, colocalisation analyses were performed in lung and cultured fibroblasts tissue (GTEx Version 8) for the corresponding gene using the coloc package[10] in R version 4.2.1. Colocalisation analyses was used to identify whether the same casual variant was associated with both IPF susceptibility and gene expression in GTEx. The posterior probability of the following models was estimated using approximate Bayes factor:

$H_0$: neither sex-specific IPF susceptibility nor gene expression have a genetic association in that region

$H_1$: only sex-specific IPF susceptibility has a genetic association in that region

$H_2$: only gene expression has a genetic association in that region

$H_3$: both sex-specific IPF susceptibility and gene expression are associated, but with different causal variants

$H_4$: both sex-specific IPF susceptibility and gene expression are associated and share a single causal variant

If the posterior probability supporting the alternative hypothesis that both sex-specific IPF susceptibility and gene expression share a single causal variant ($H_4$) was greater than 80% then we concluded that the sex-specific IPF and gene expression signal colocalised.

#### Polygenic risk score analysis

Two main polygenic risk score analyses (PRS) were performed, 'standard PRS' and 'sex-specific PRS'. The 'base data' were derived from the meta-analyses of the US, Colorado, UK, UUS and Genentech datasets[2] (4,096 cases & 20,433 controls) with the association effect sizes from the previously published combined-sex IPF GWAS meta-analysis used for the 'standard PRS', and association effect sizes from new sex-specific IPF GWAS meta-analyses of the same five datasets used for the 'sex-specific PRS' **(Figure 1)**. The 'target dataset' was the CleanUP-UCD study, which comprised 2,297 males (372 cases & 1,925 controls) and 623 females (93 cases & 530 controls). Bi-allelic autosomal variants that were well imputed (imputation $R^2 \geq 0.5$), had a MAF ≥ 0.01 and did not depart from Hardy-Weinberg Equilibrium ($P \geq 1\times10^{-6}$) were retained in the base data. Ambiguous SNPs were excluded. Only variants available in both the base data and the target dataset were included in the analyses.

For the 'standard PRS' we first constructed the 19-variant PRS using the effect sizes from the base data[2]. We then tested the predictive accuracy of this PRS in males and females separately in the target data. Using the same base data, we then created multiple PRSs for a range of *p*-value thresholds ($P_T$) using PRSice v2.3.5[11] and the PRS threshold with the most significant *p*-value association in the target data was selected as the best-performing PRS.

For the 'sex-specific PRS' we used sex-specific GWAS results derived from the base data and PRSice to create multiple PRSs in males and females separately. The PRS threshold with the most significant *p*-value association in the two target datasets separately (CleanUP-UCD male and CleanUP-UCD female) were selected as the best-performing PRS.

For the best performing PRS in both the 'standard PRS' and 'sex-specific PRS' analyses, we also estimated the AUC to examine its predictive accuracy. For the 'sex-specific' analysis we tested whether the predictive accuracy of a PRS constructed in males using male-specific effect sizes was statistically significantly different to a PRS constructed in females using female-specific effect sizes, using DeLong's test.

For all PRS analyses, linkage disequilibrium (LD) was accounted for using clumping ($R^2$> 0.1 across 250Kb window).

### Supplementary Tables

**Table S1: Sentinel variants that reach $P$<1×10$^{-6}$ and posterior probability of replication (MAMBA) >90%** [Excel spreadsheet]

**Table S2: Annotation and eQTL results for variants in 95% credible set of IPF SNP-by-sex signals** [Excel spreadsheet]

**Table S3: Results of colocalisation analysis between rs62040020 and lung tissue and cultured fibroblasts using Coloc (female and male specific results)** [Excel spreadsheet]

**Table S4: SNP-by-sex interaction meta-analysis results for previously reported IPF susceptibility SNPs** [Excel spreadsheet]

**Table S5: Polygenic risk score analyses results i) 'standard PRS' analysis ii) 'sex-specific PRS'** analysis [Excel spreadsheet]

#### Supplementary Figures

**Figure S1:** Quantile-quantile (QQ) plot for SNP-by-sex interaction results.

QQ plot for the genome-wide SNP-by-sex interaction results with expected $-\log_{10}$(*P* value) on the x-axis and the observed $-\log_{10}$(*P* value) on the y-axis. (Inflation factor λ = 1.02)

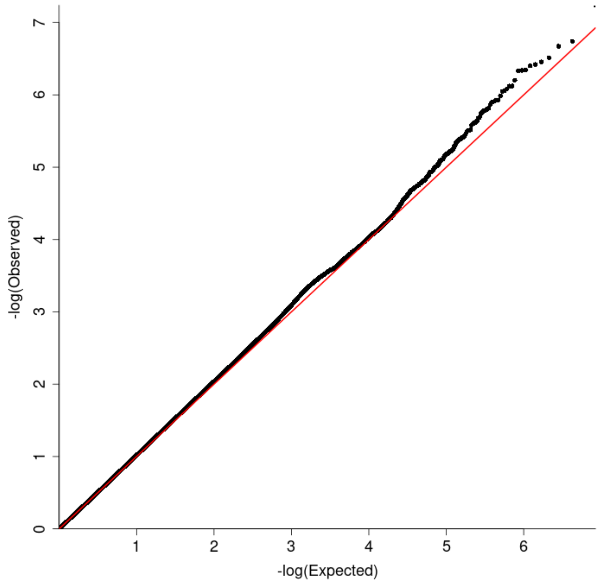

**Figure S2:** Forest plot for of male only results by IPF study and meta-analysed male results for **a)** rs62040020, **b)** rs1756167317 and **c)** rs1663078846.
OR = odds ratio and CI = confidence interval

**a)**

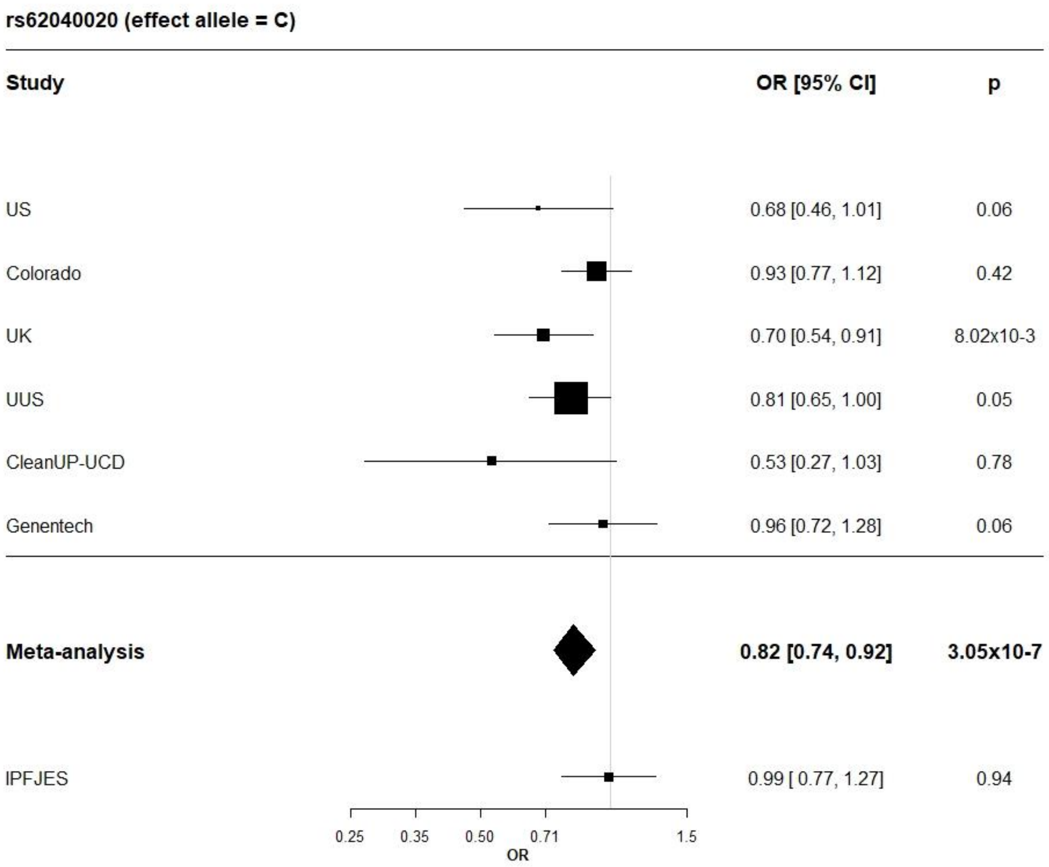

b)

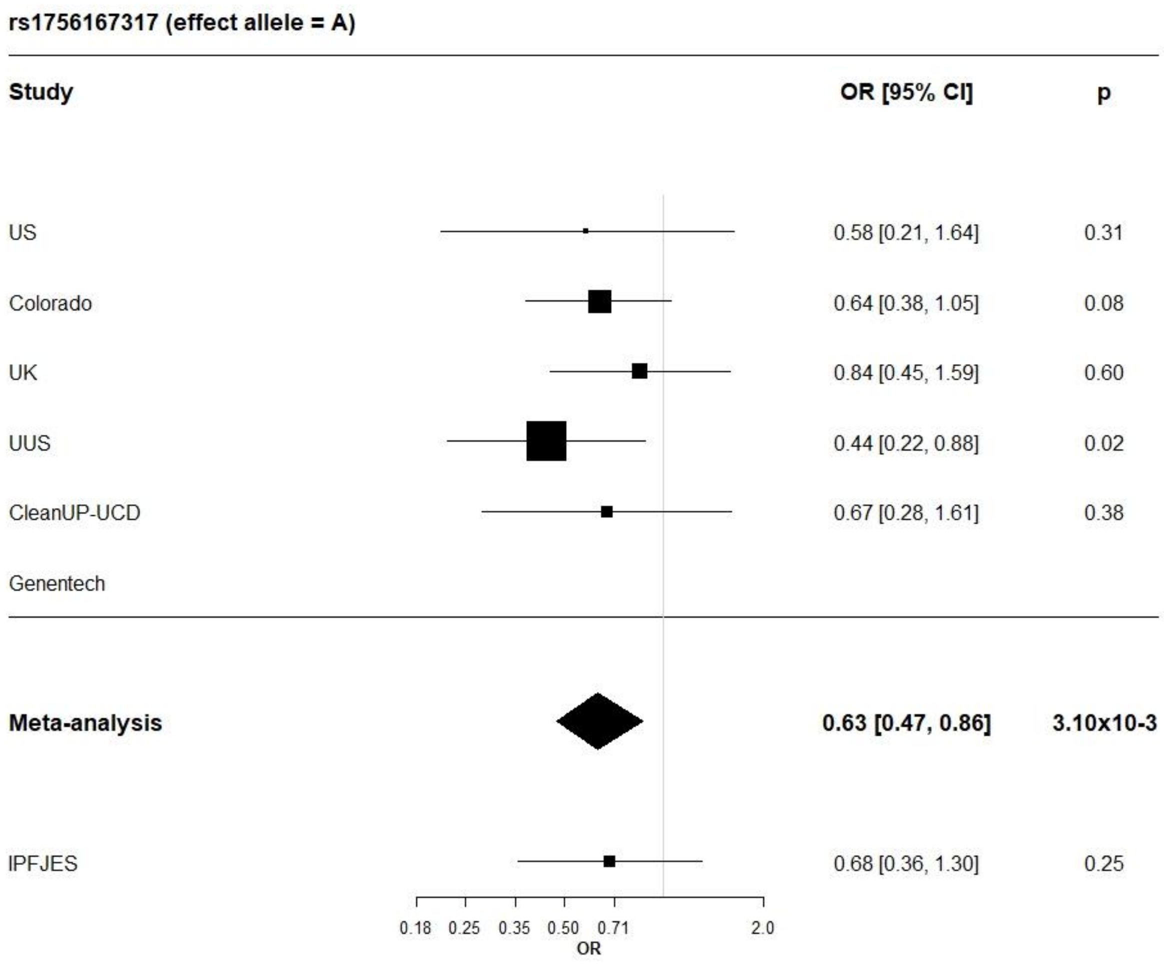
rs1756167317 (effect allele = A)
Study
OR [95% CI]
p
US
0.58 [0.21, 1.64]
0.31
Colorado
0.64 [0.38, 1.05]
0.08
UK
0.84 [0.45, 1.59]
0.60
UUS
0.44 [0.22, 0.88]
0.02
CleanUP-UCD
0.67 [0.28, 1.61]
0.38
Genentech
Meta-analysis
0.63 [0.47, 0.86]
3.10x10-3
IPFJES
0.68 [0.36, 1.30]
0.25
0.18
0.25
0.35
0.50
0.71
2.0
OR

c)

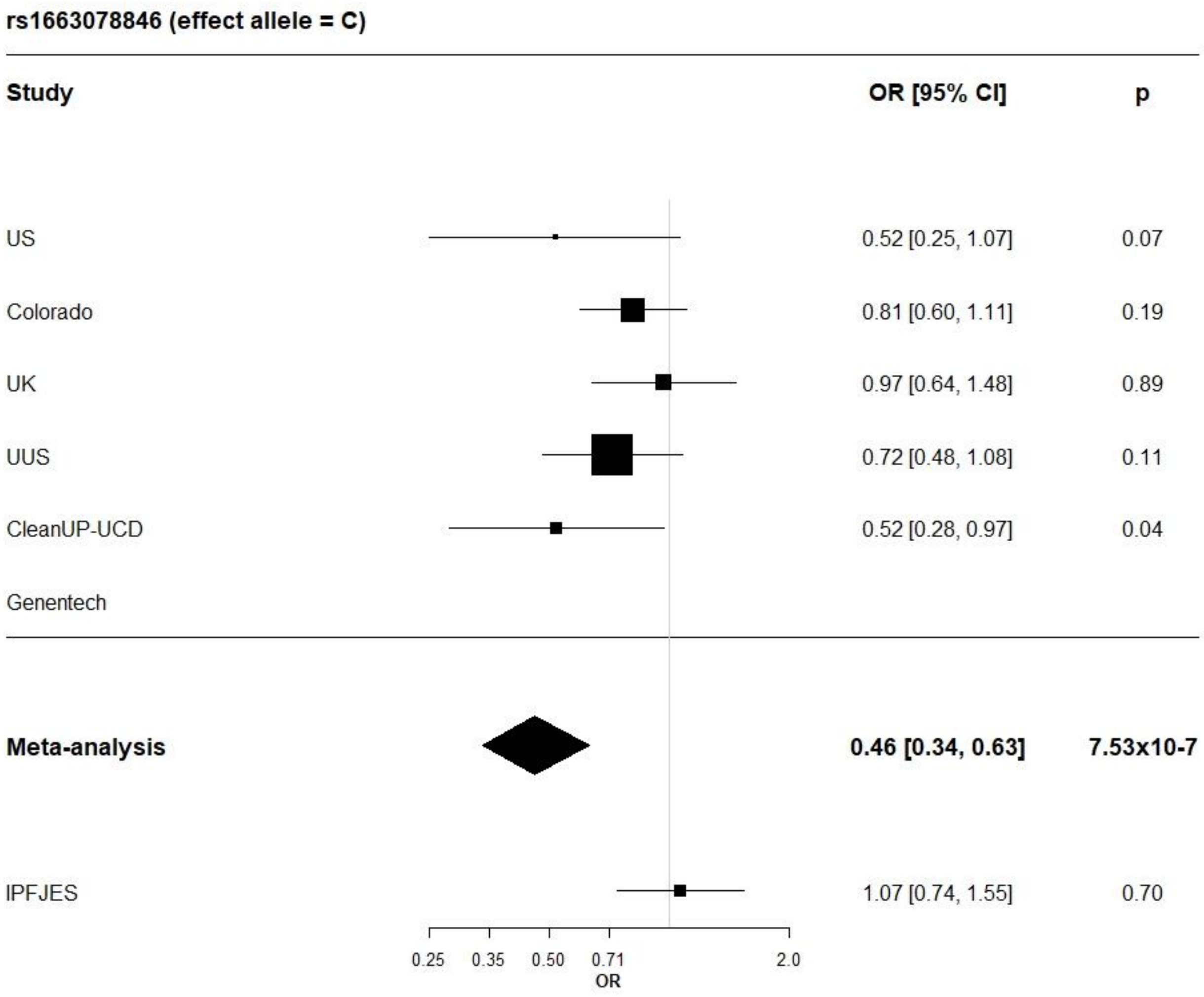
rs1663078846 (effect allele = C)
Study
OR [95% CI]
p
US
0.52 [0.25, 1.07]
0.07
Colorado
0.81 [0.60, 1.11]
0.19
UK
0.97 [0.64, 1.48]
0.89
UUS
0.72 [0.48, 1.08]
0.11
CleanUP-UCD
0.52 [0.28, 0.97]
0.04
Genentech
Meta-analysis
0.46 [0.34, 0.63]
7.53x10-7
IPFJES
1.07 [0.74, 1.55]
0.70
0.25
0.35
0.50
0.71
2.0
OR

**Figure S3:** Sex-stratified GWAS results vs eQTL results for rs62040020 (build 38 - chr16: 1697584) for a) female specific GWAS and b) male specific GWAS. The chromosomal position is on the x-axis and the -log(p-value) for each genetic variant is on the y-axis. On the y-axis above the x-axis is the -log(p-value) from the sex-stratified GWAS results (female or male only results) and on the y-axis below the x-axis is the -log(p-value) from the eQTL database. The sentenal variant (rs62040020) is coloured blue and all other variants are coloured by their linkage disequlibrium ($r^2$) with rs62040020.

**a) female specific GWAS**

***i)*** ***HAGH*** **- Lung**

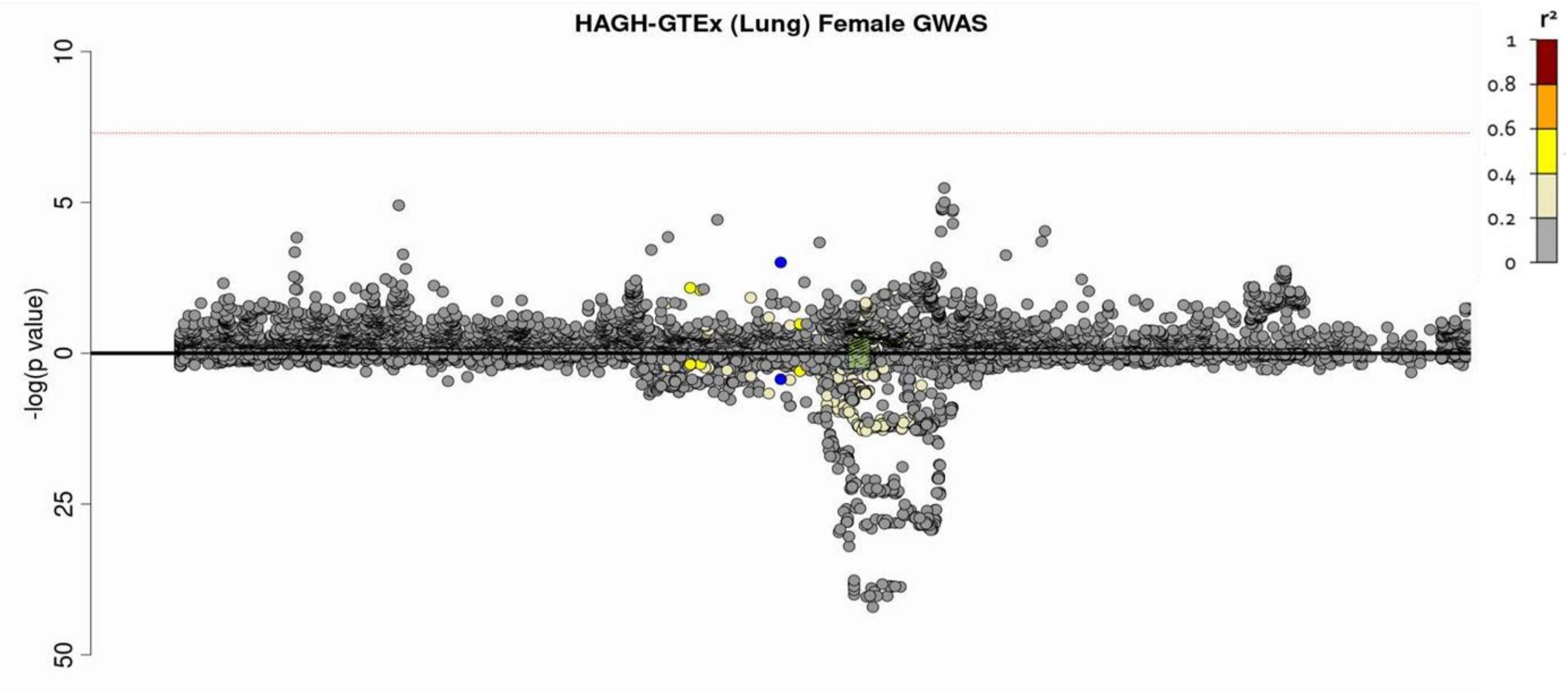

***ii)*** ***MEIOB*** **- Lung**

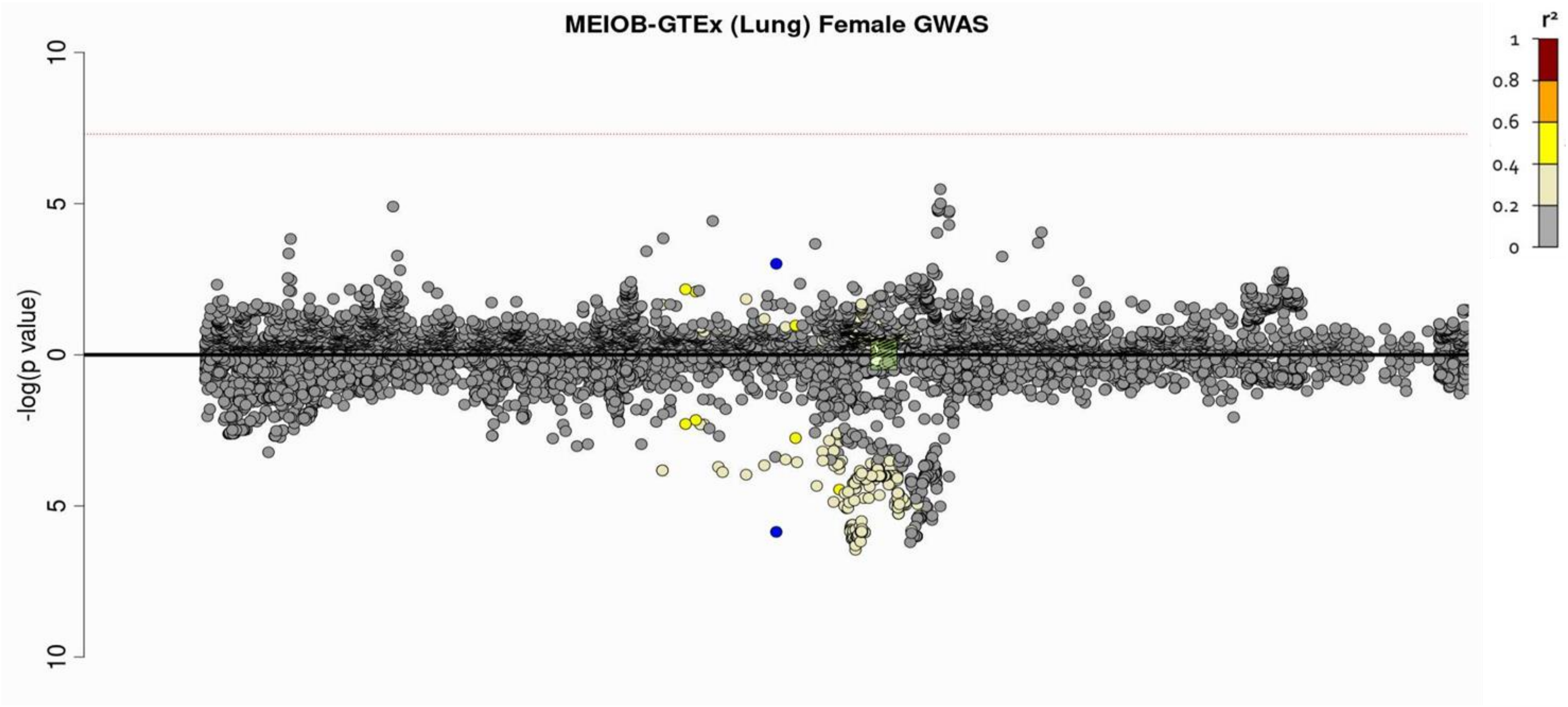

***iii)*** ***FAHD1*** **- Lung**

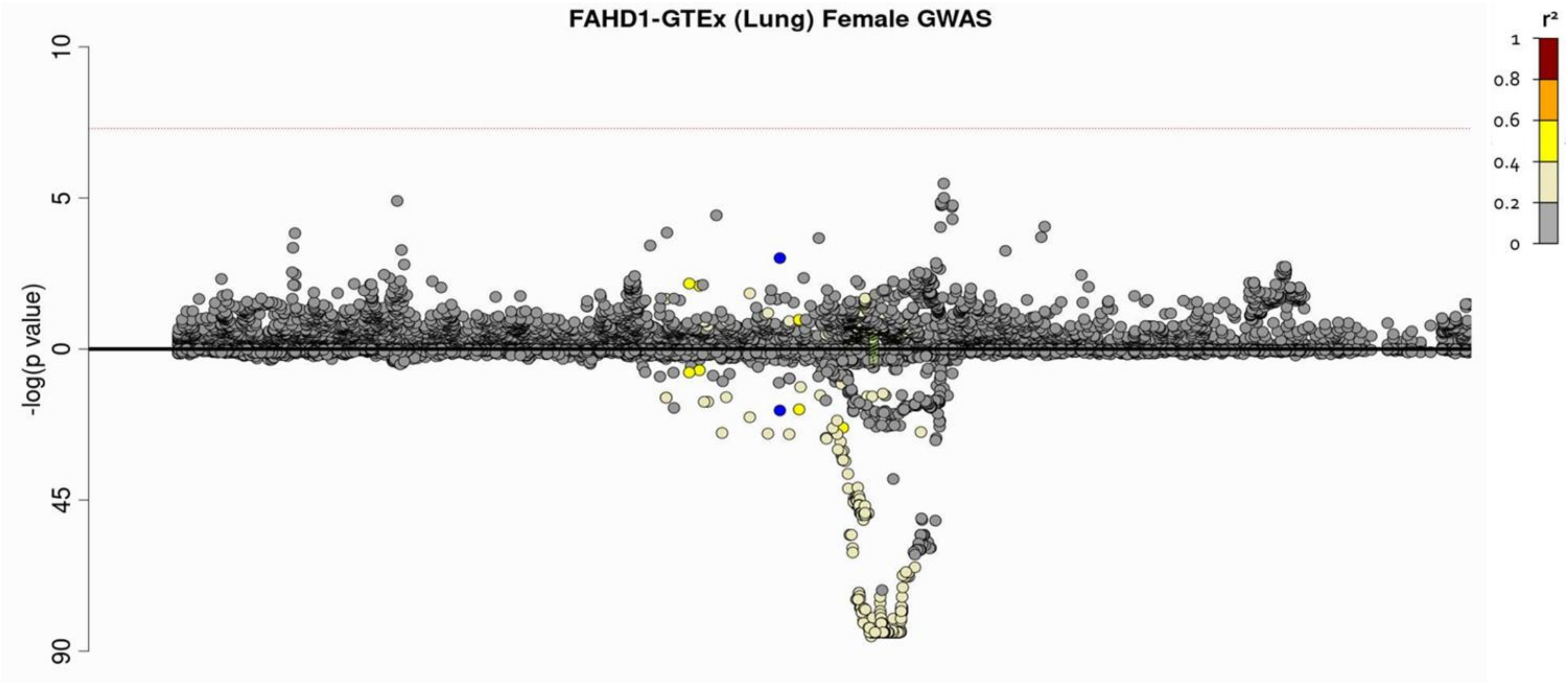

*iv)* *HAGH* **– Cultured Fibroblasts**

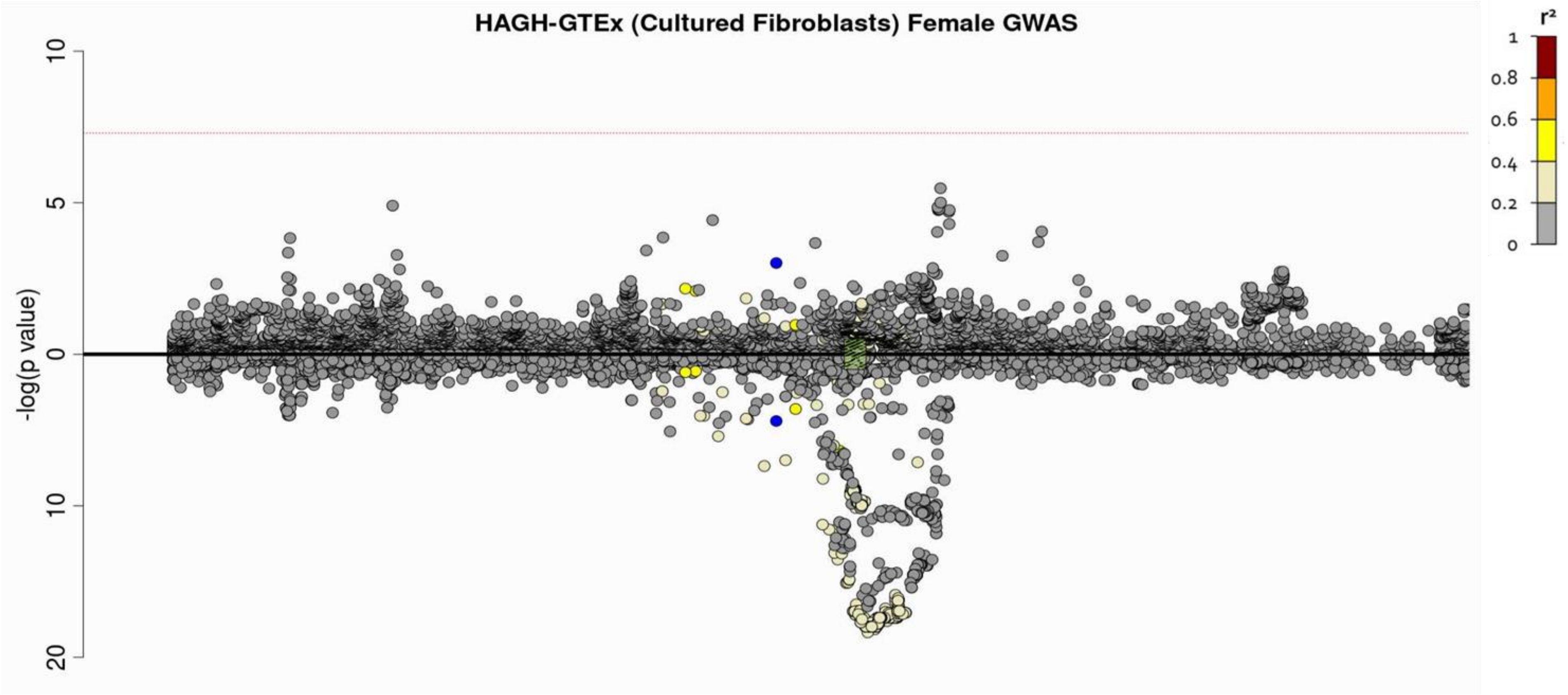

***v)*** ***MEIOB* – Cultured Fibroblasts**

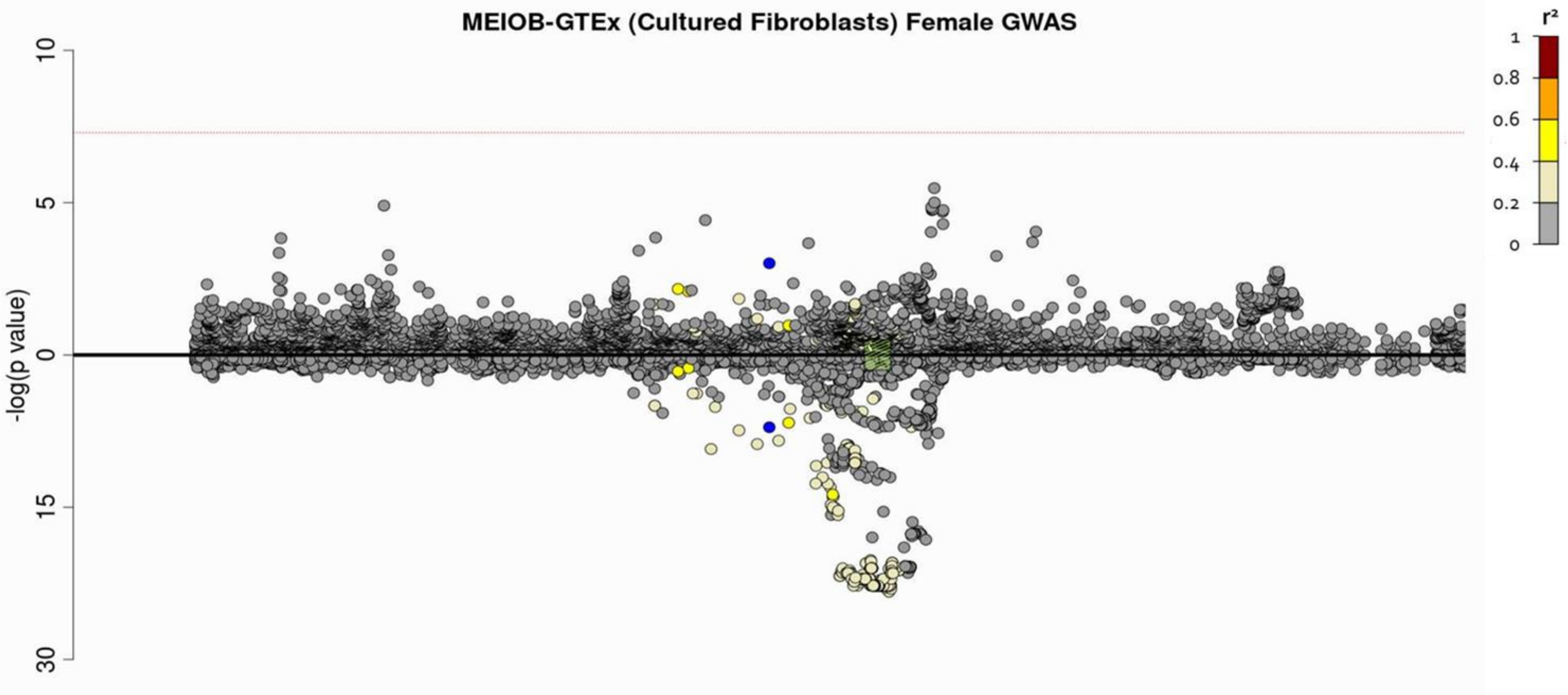

***vi)*** ***FAHD1*** **– Cultured fibroblasts**

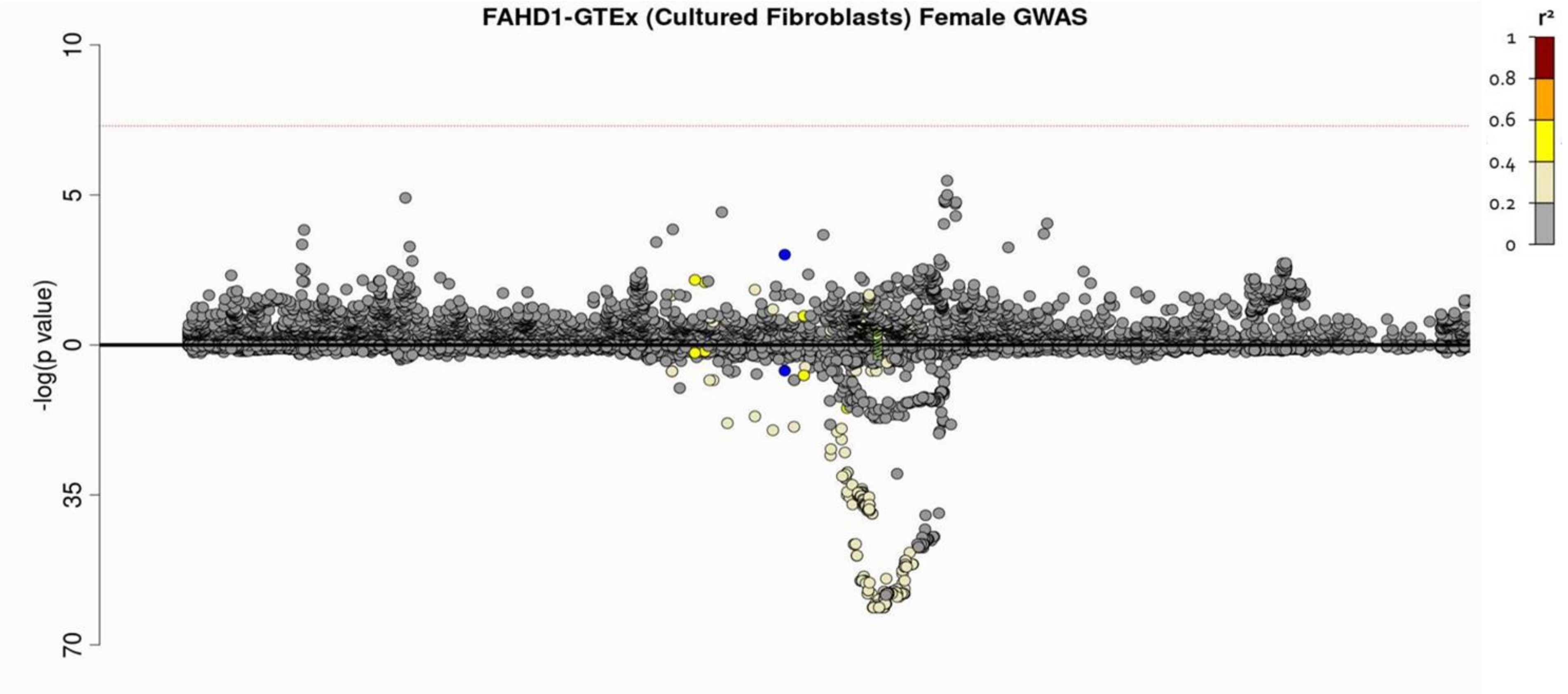

***vii)*** **MRPS34 – Cultured fibroblasts**

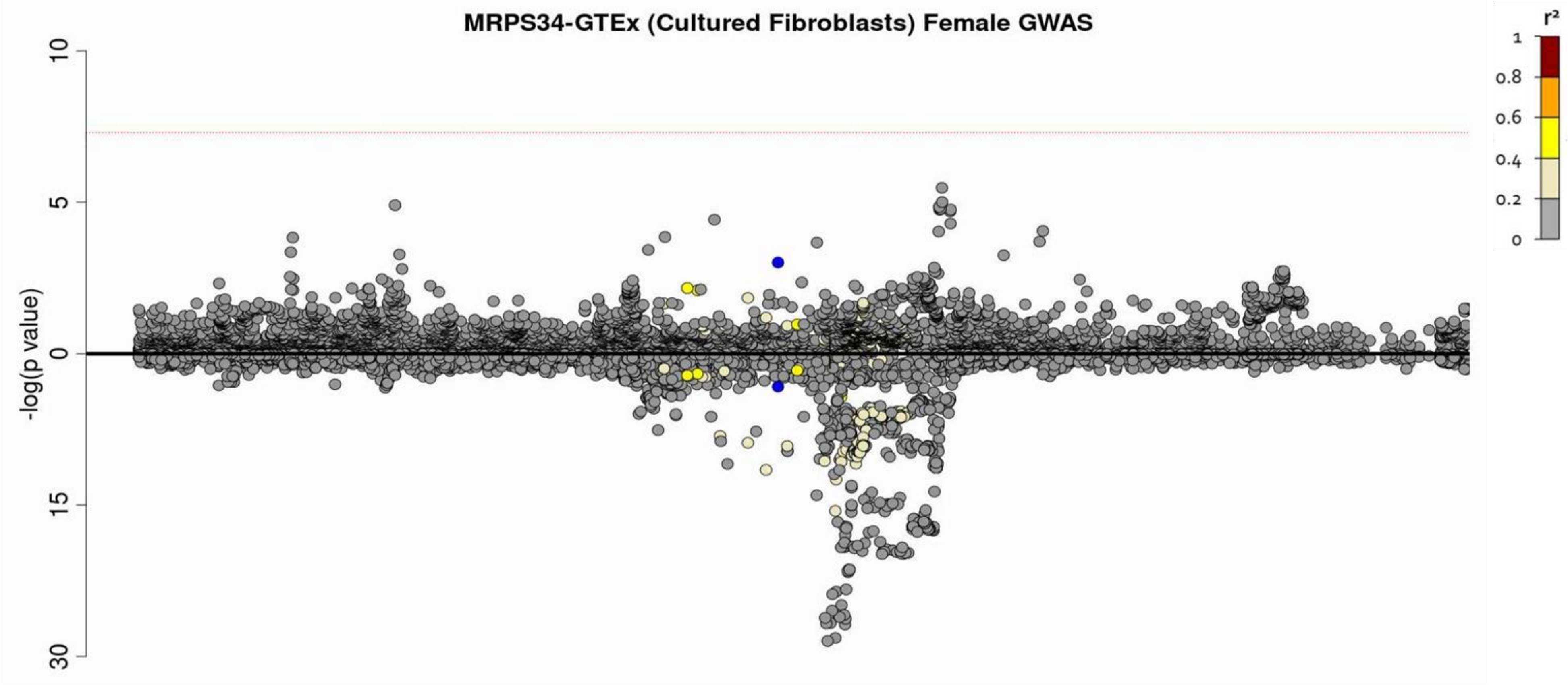

***viii)*** **NUBP2 – Cultured fibroblasts**

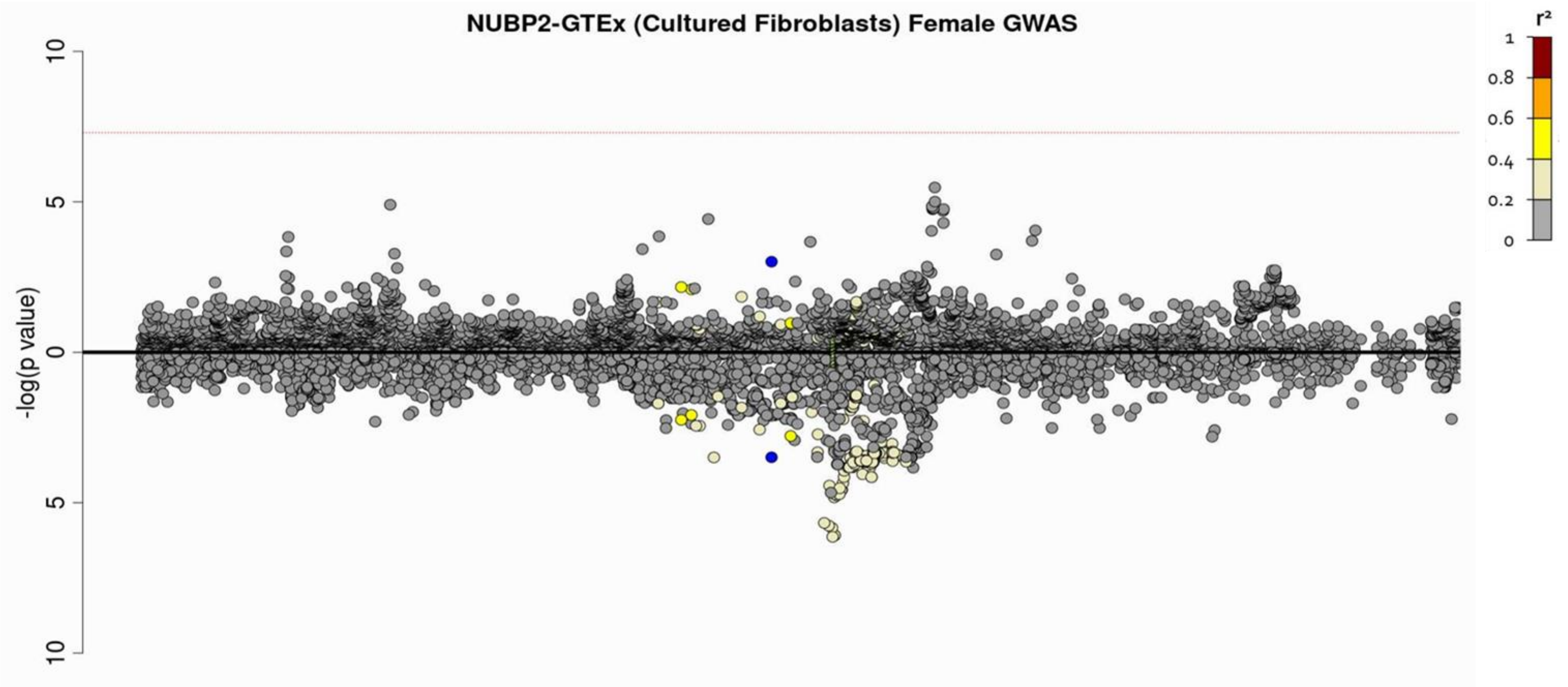

**b) male specific GWAS**

***i)*** ***HAGH*** **- Lung**

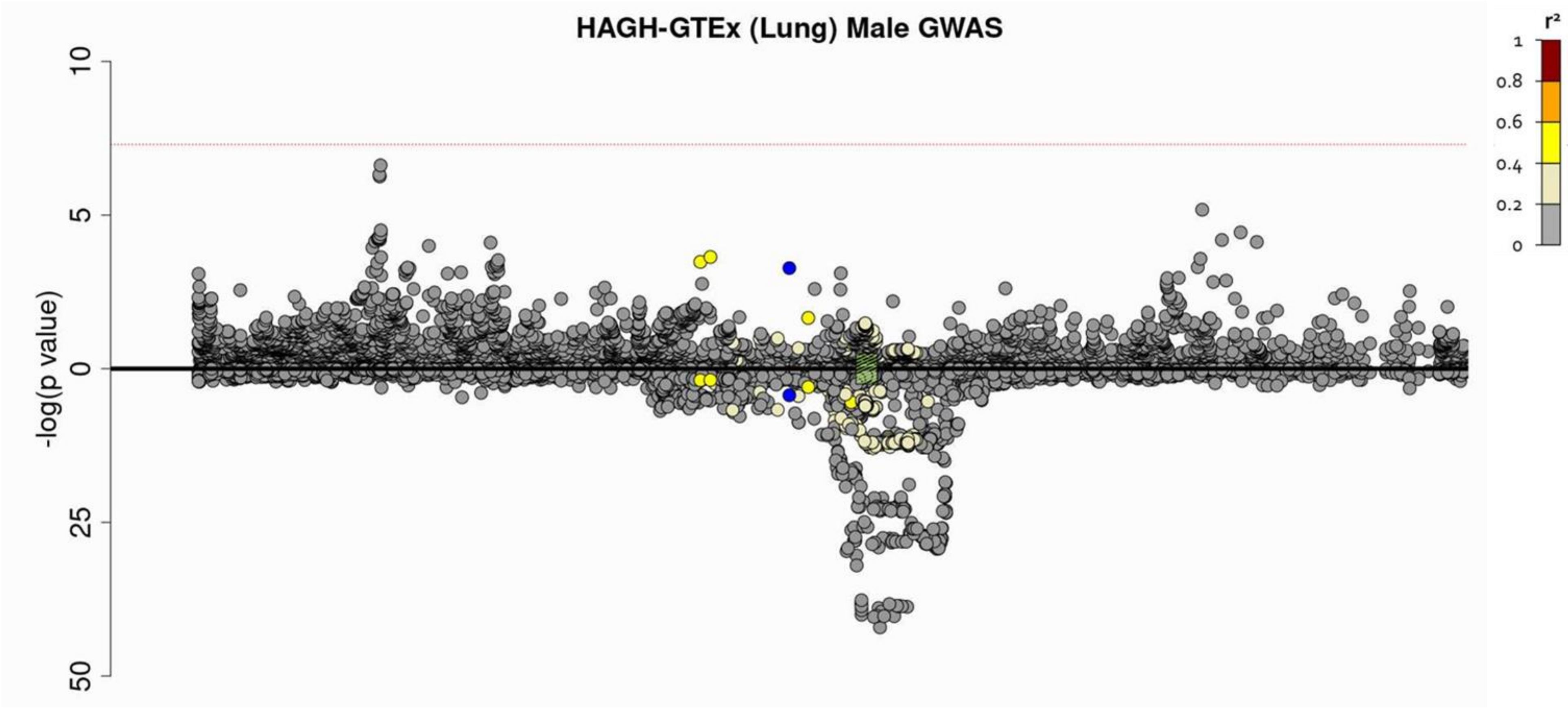

***ii)*** ***MEIOB*** **- Lung**

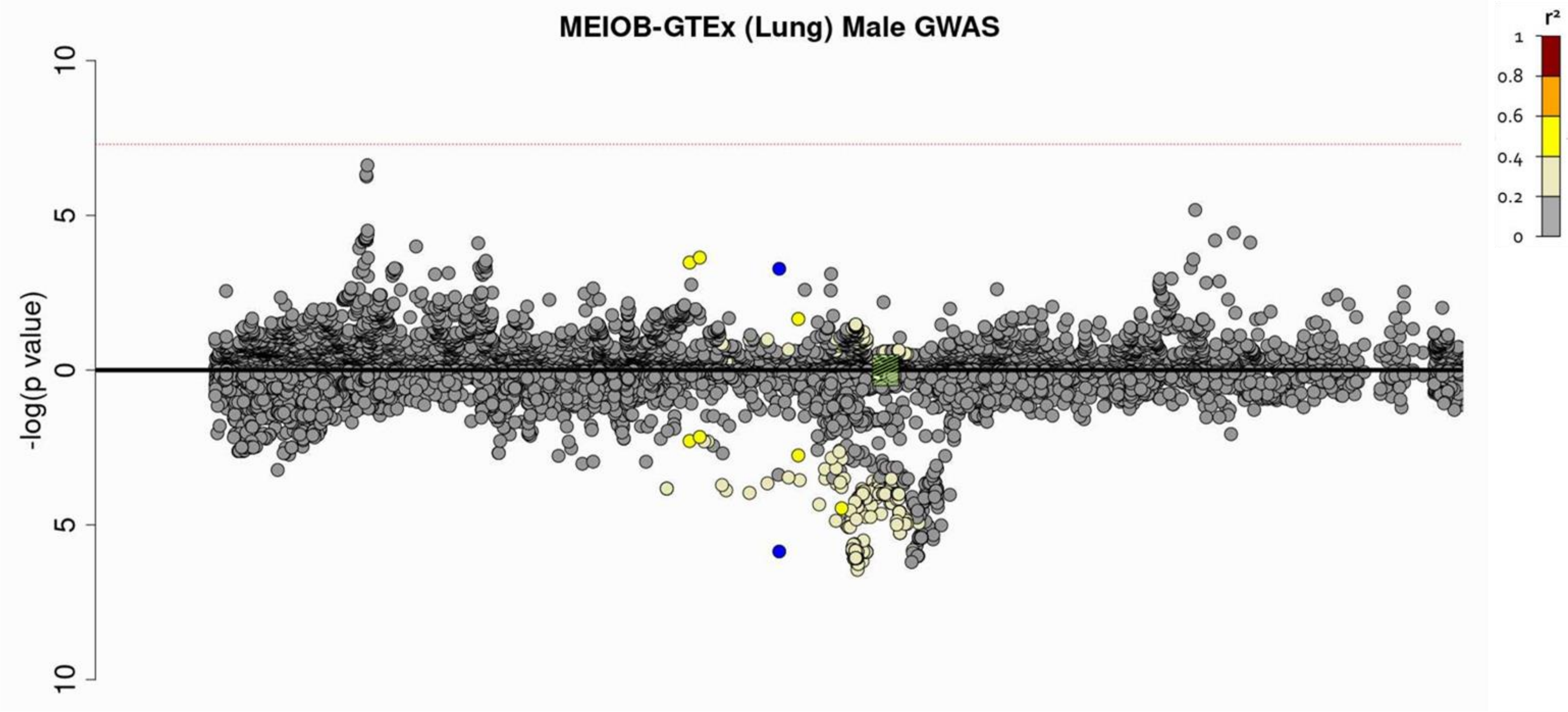

***iii)*** ***FAHD1*** **- Lung**

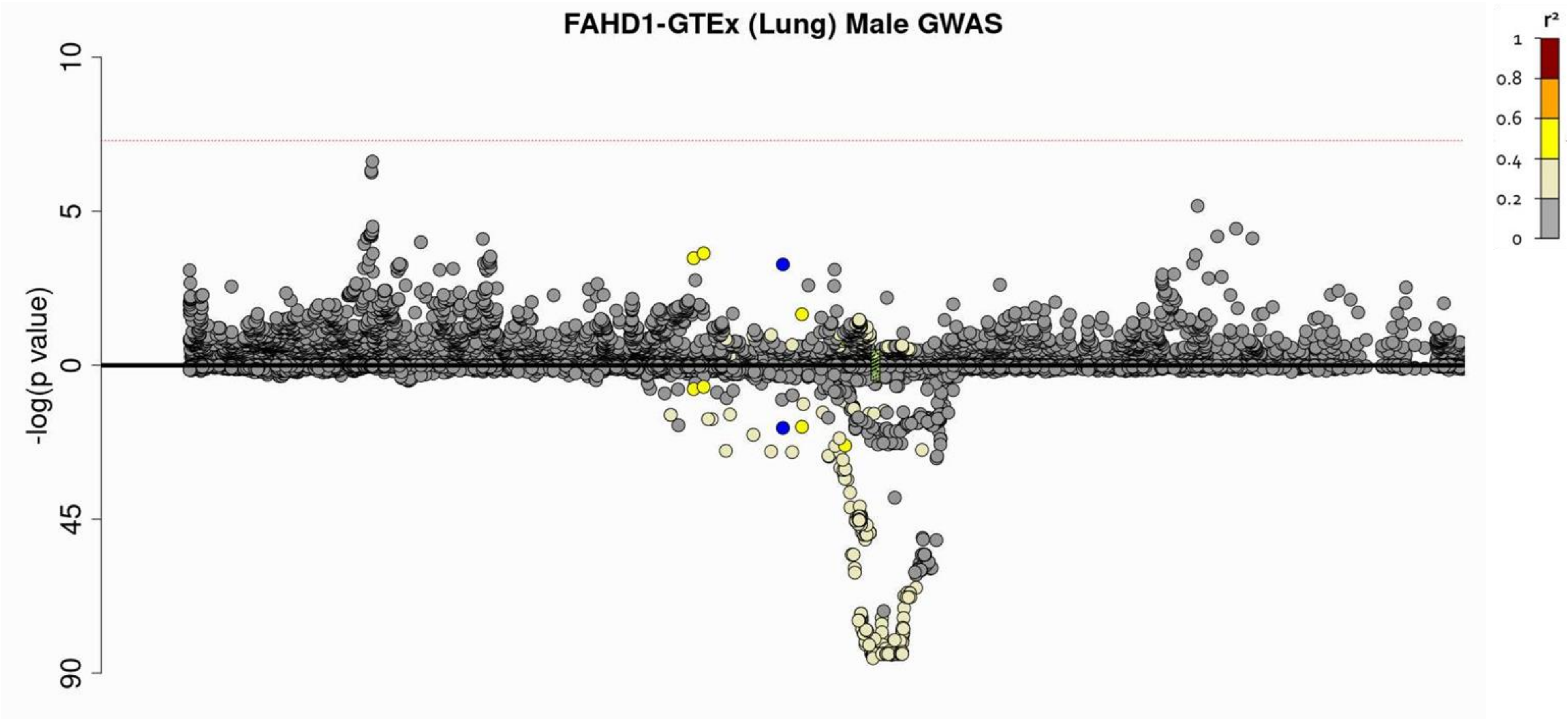

***iv)*** ***HAGH*** **– Cultured Fibroblasts**

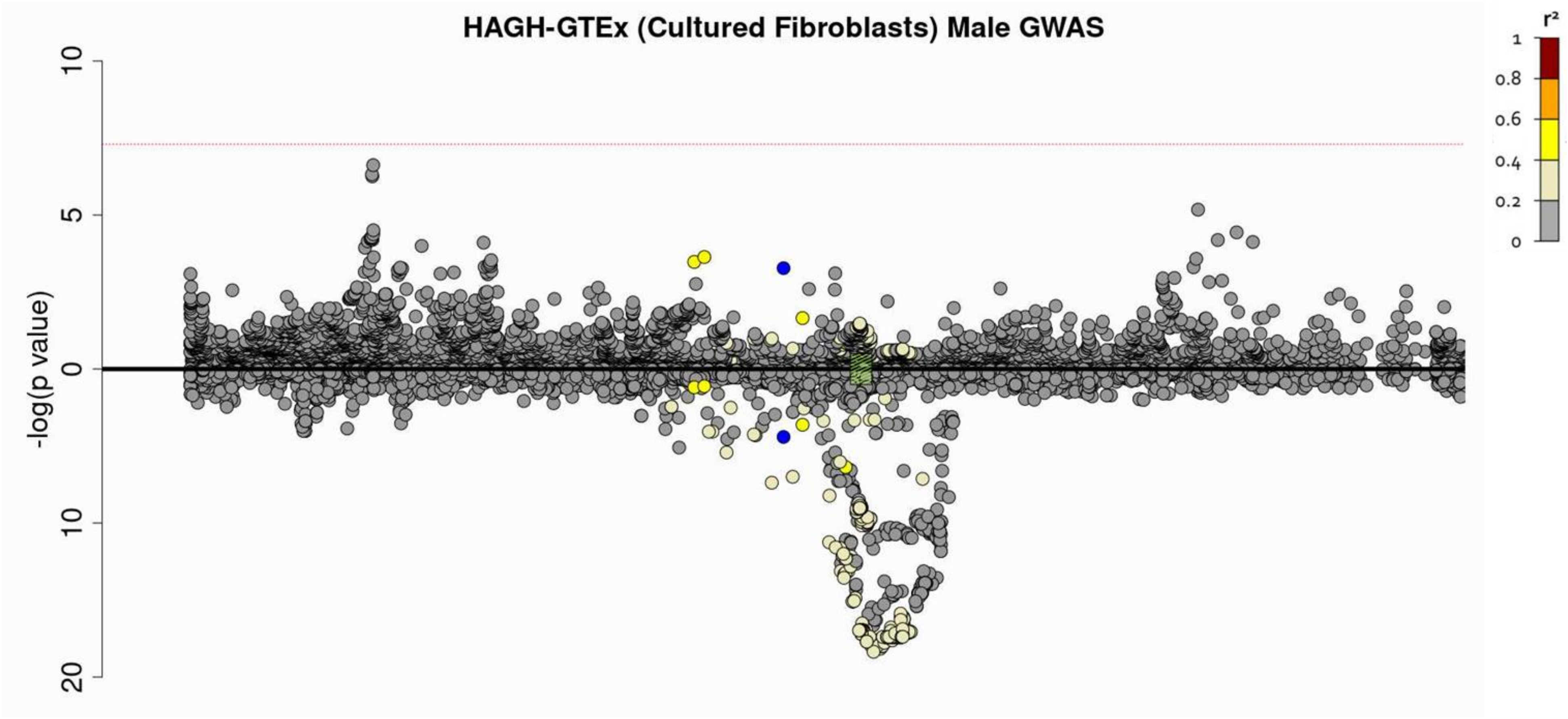

***v)*** ***MEIOB*** **– Cultured Fibroblasts**

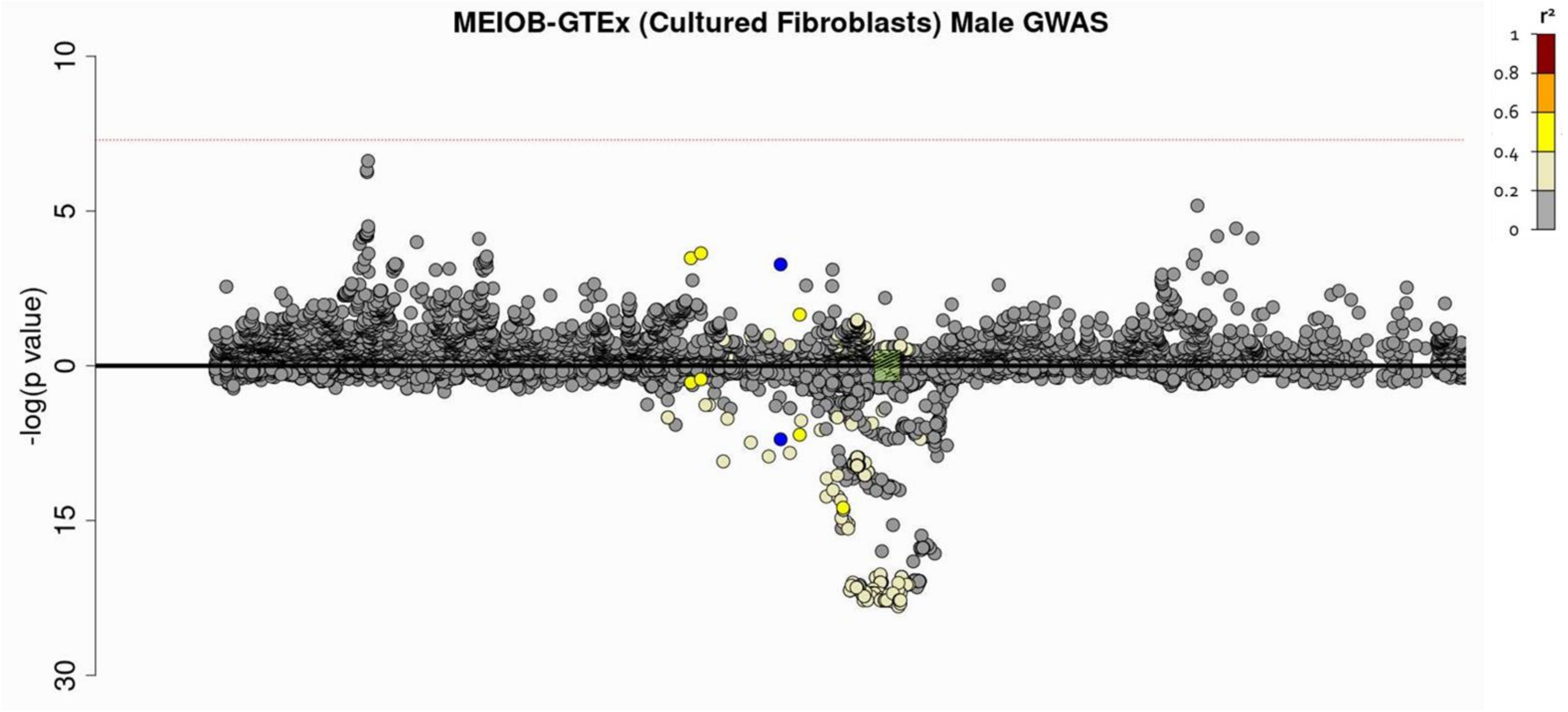

***vi)*** ***FAHD1*** **– Cultured Fibroblasts**

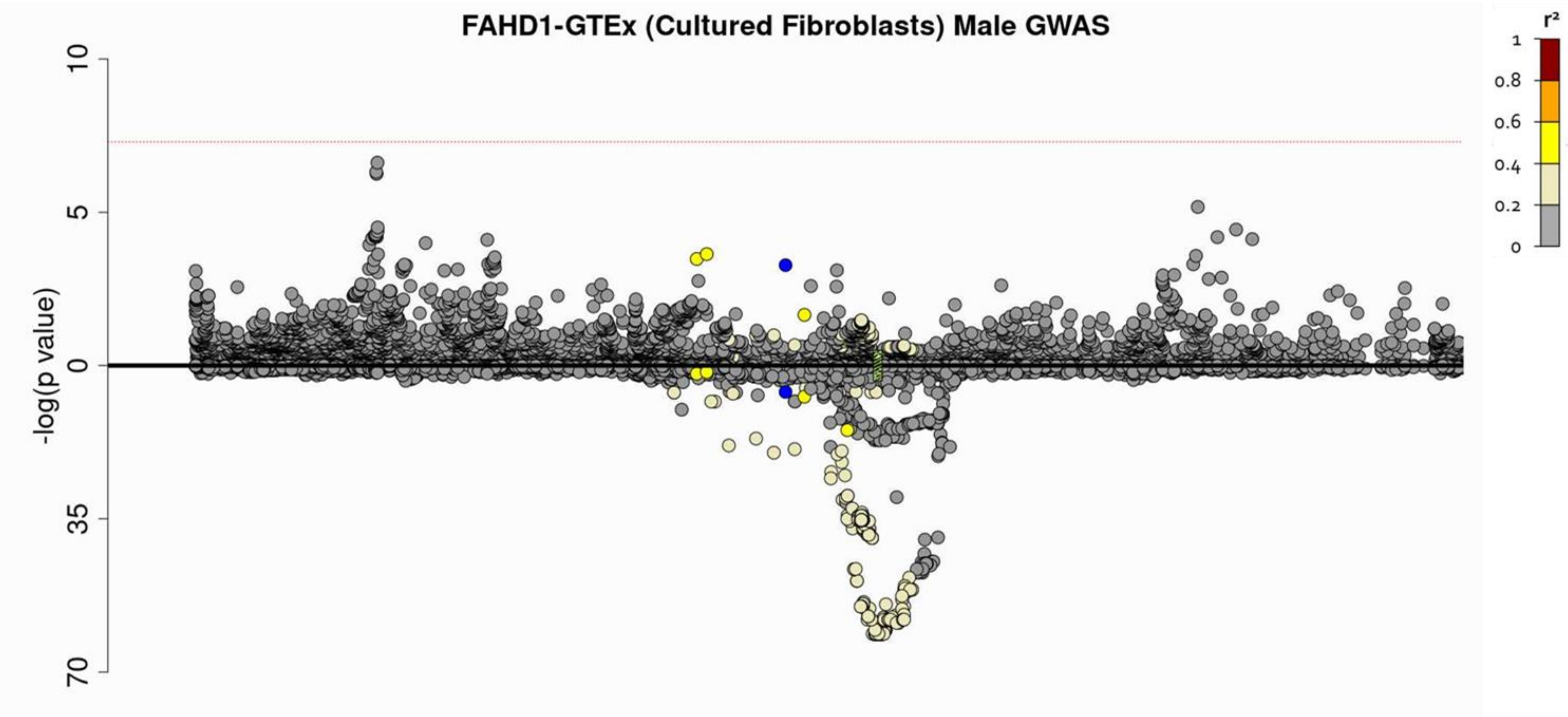

***vii) MRPS34* – Cultured Fibroblasts**

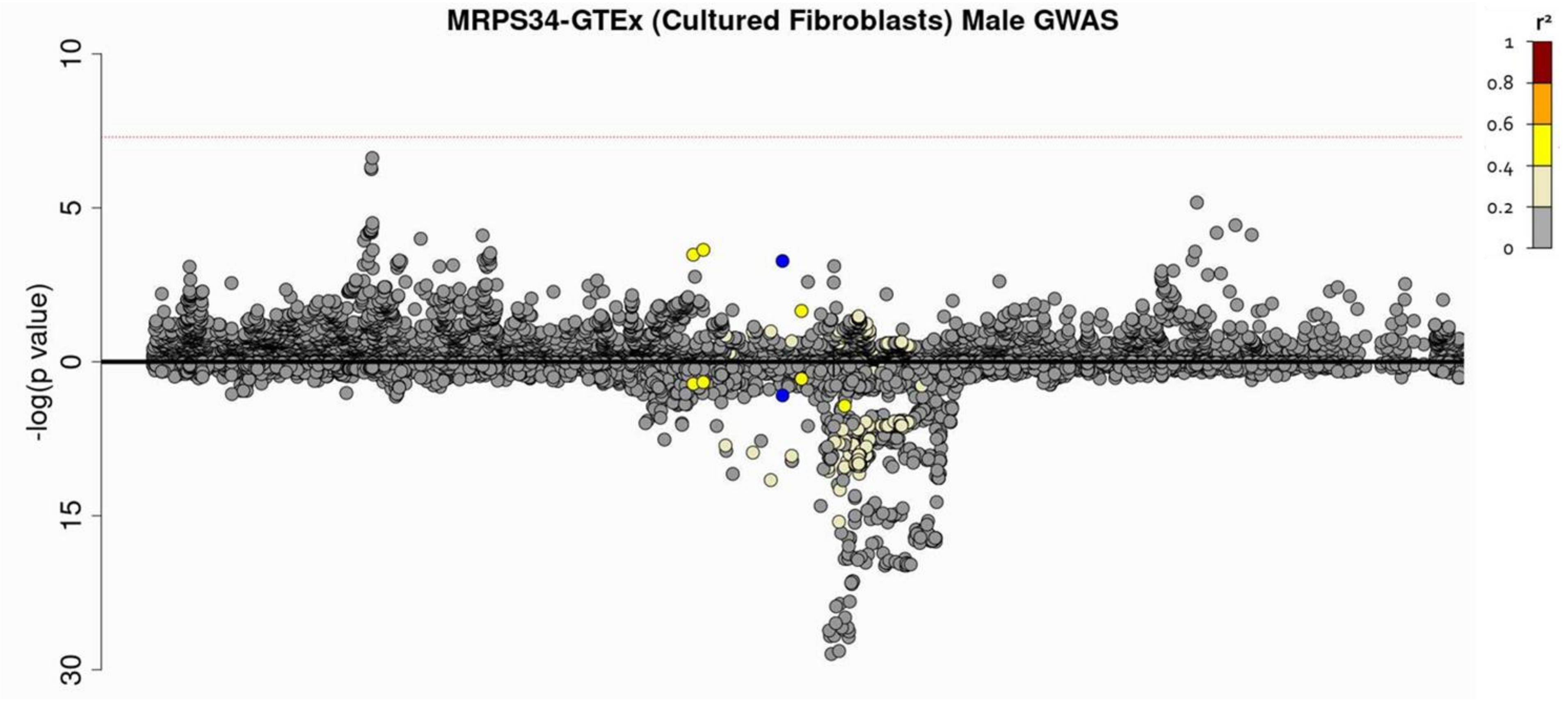

***viii)*** ***NUBP2* – Cultured Fibroblasts**

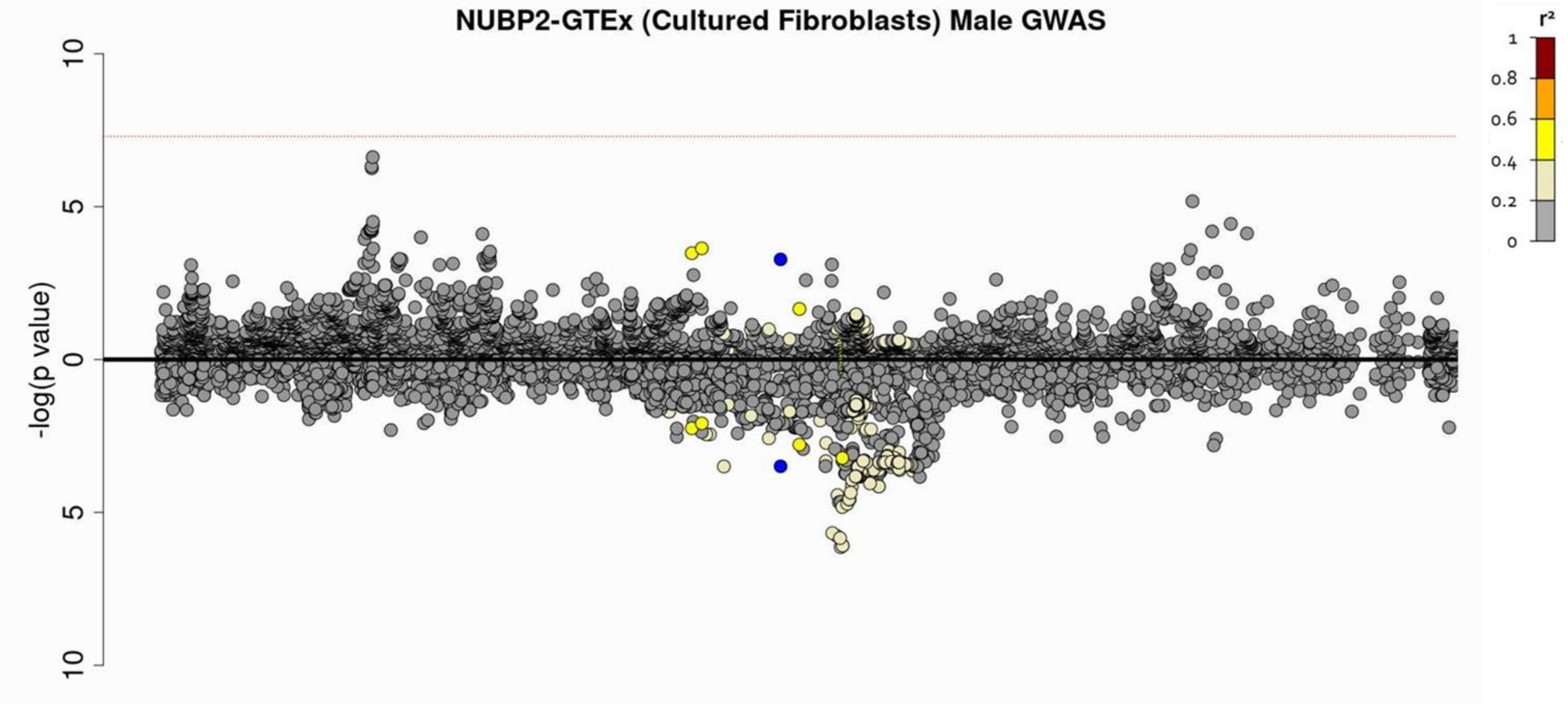

**Figure S4:** Forest plots showing SNP-sex interaction odds ratio by study and the meta-analysed result for rs2076295 (*DSP*)
OR = odds ratio and CI = confidence interval

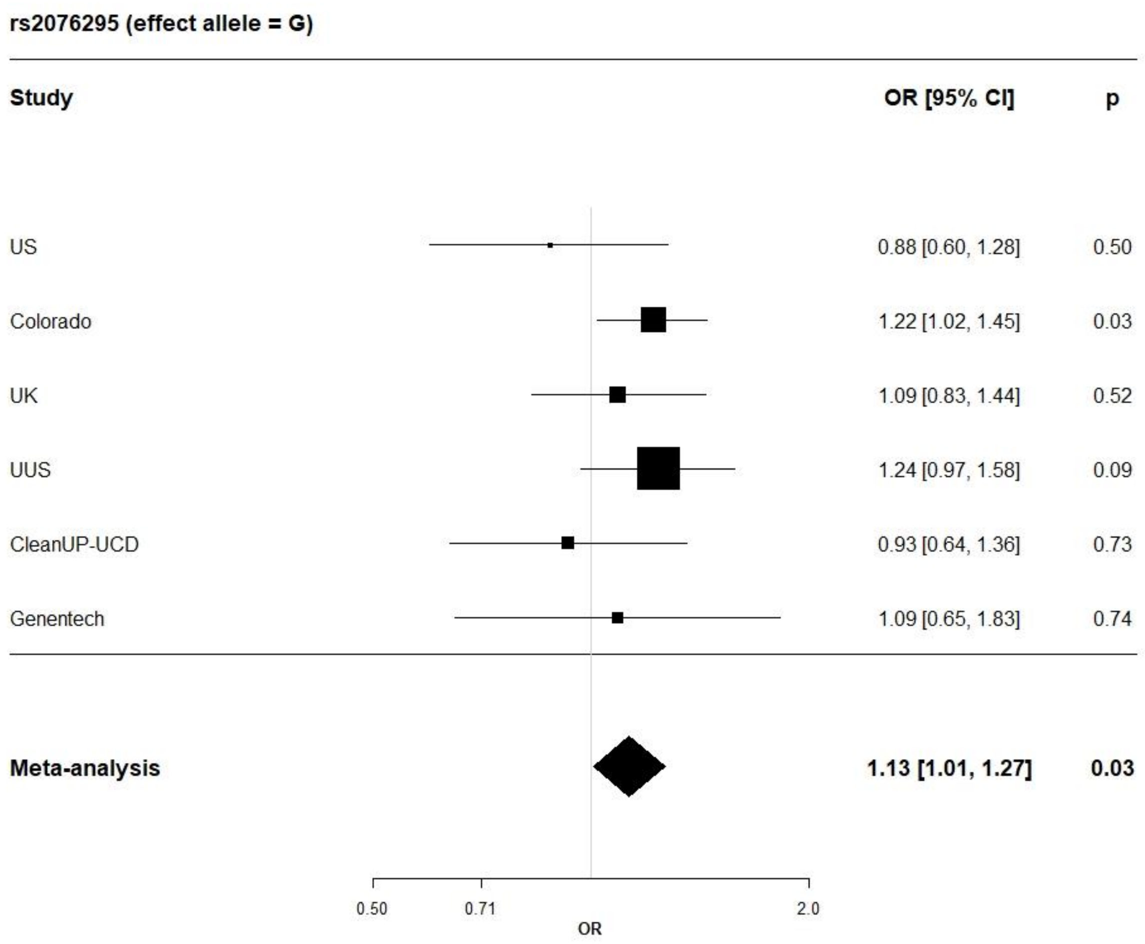

**Figure S5:** Strength of association for polygenic risk score when using different *p*-value threshold (Pt) for a) beta estimates from combined GWAS and target dataset contain only males from CleanUp-UCD study, b) beta estimates from combined GWAS and target dataset contain only females from CleanUp-UCD study, c) beta estimates from male-specific GWAS and target dataset contain only males from CleanUp-UCD study and d) beta estimates from female-specific GWAS and target dataset contain only females from CleanUp-UCD study.
*Note: The plots are zoomed in, so the the y-axis on the plots do not start at 0 in order to help with visualisation.*

a) beta estimates from combined GWAS and target dataset contain only males from CleanUp-UCD study

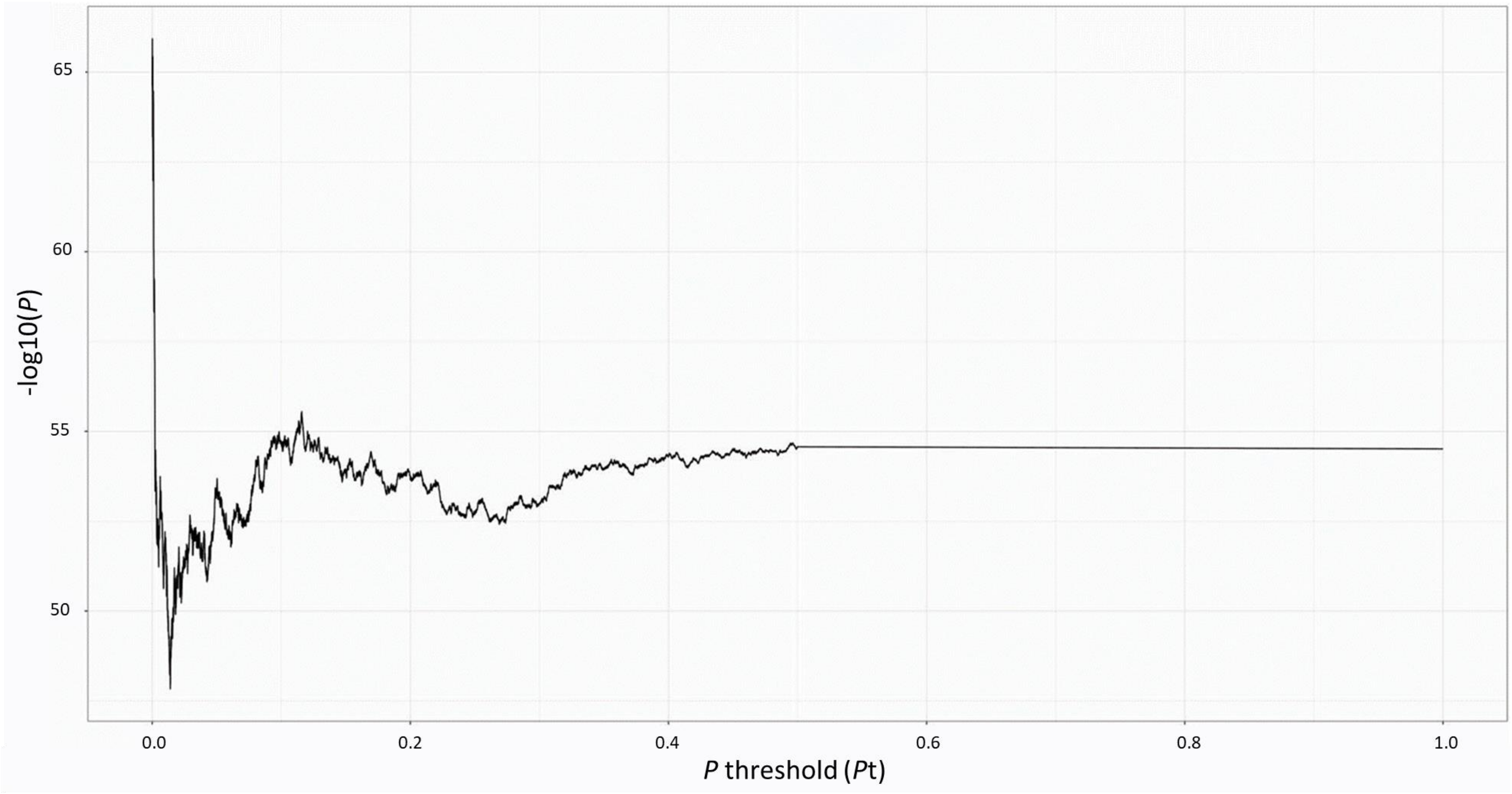

b) beta estimates from combined GWAS and target dataset contain only females from CleanUp-UCD study

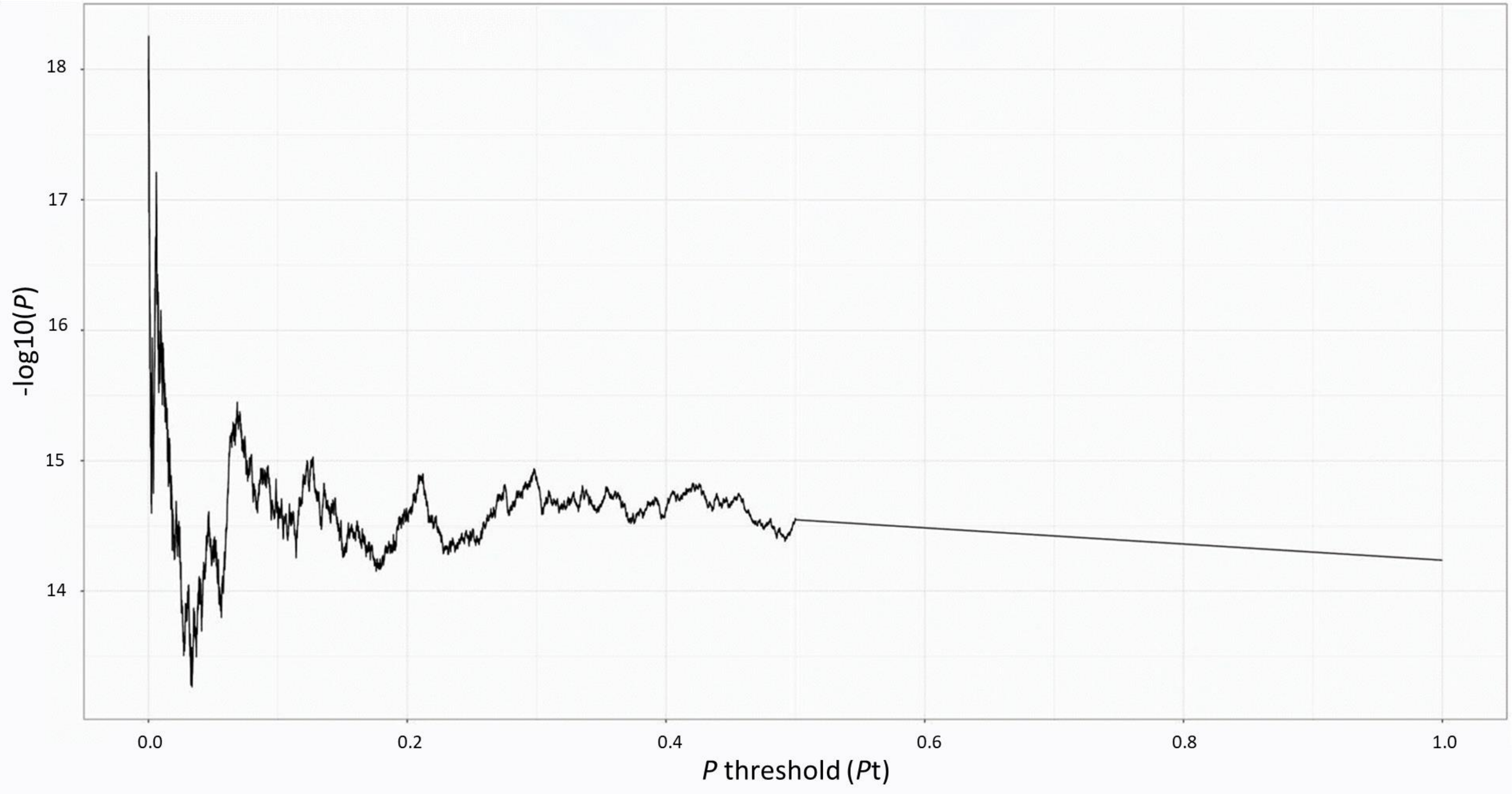

c) beta estimates from male-specific GWAS and target dataset contain only males from CleanUp-UCD study

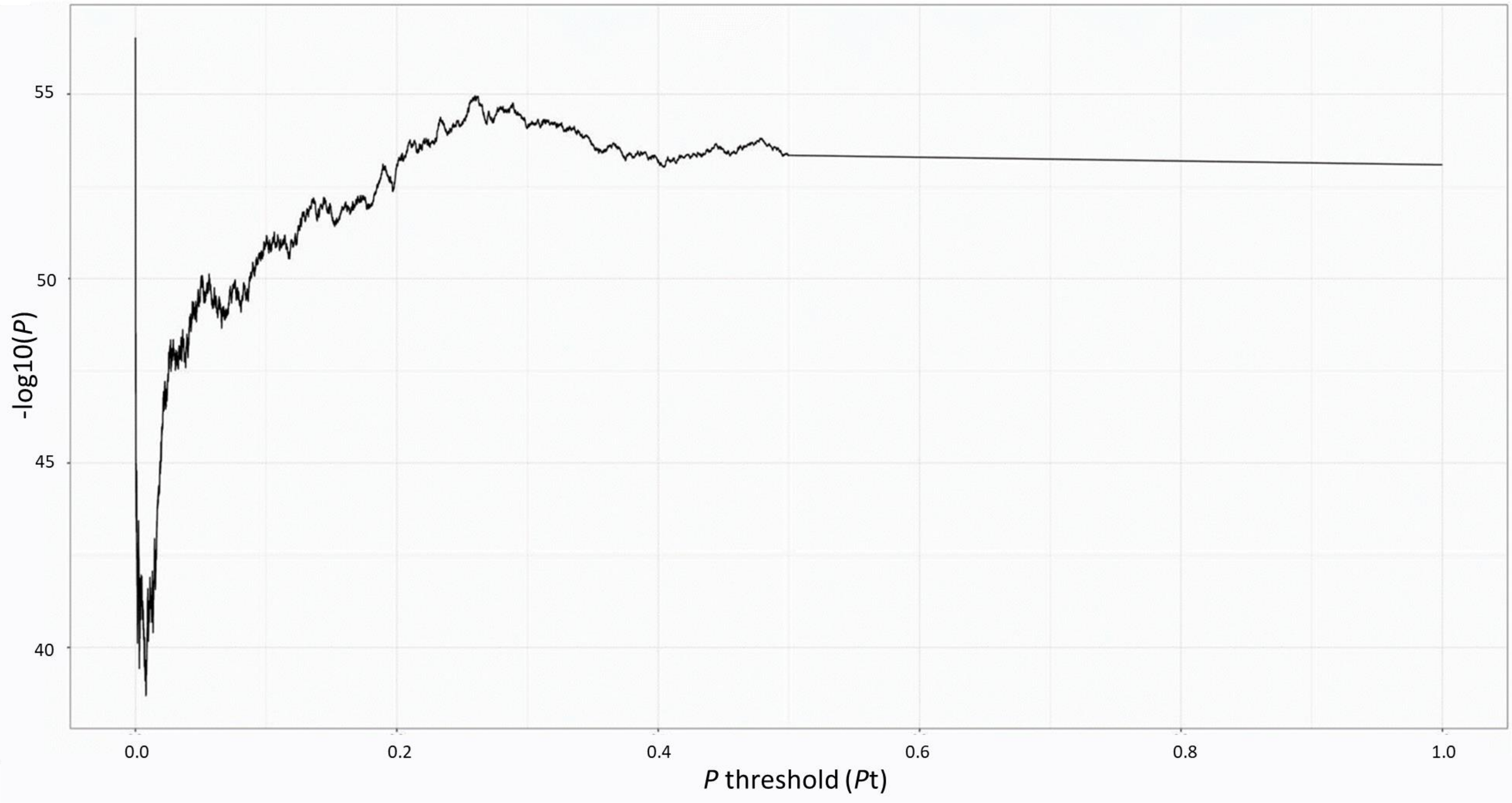

d) beta estimates from female-specific GWAS and target dataset contain only females from CleanUp-UCD study

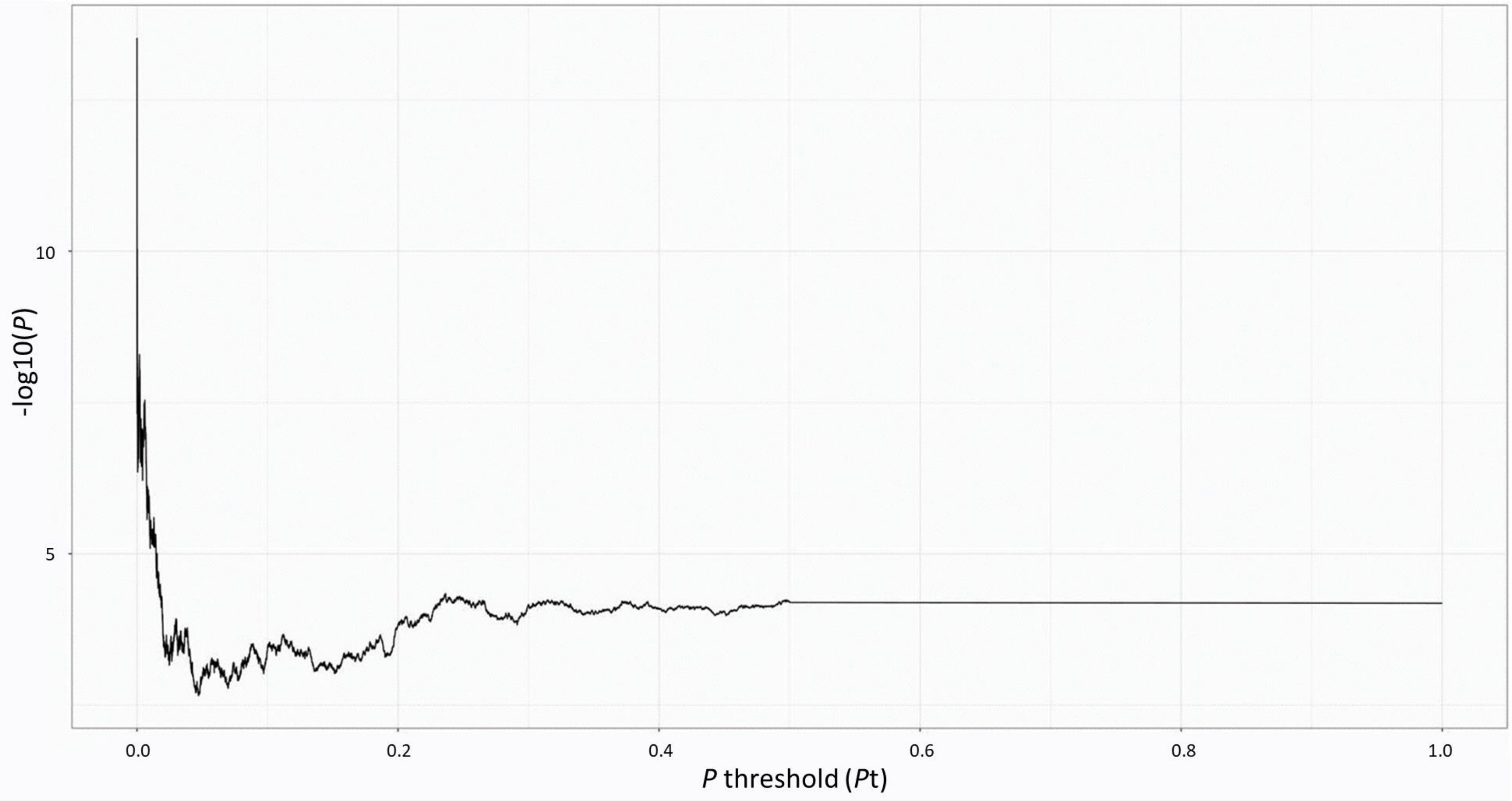
